# Biological age measured by DNA methylation clocks and neurodegenerative diseases: A systematic review and meta-analysis

**DOI:** 10.64898/2026.10.21.26365867

**Authors:** Zaylea Kua, Jian Hua Tay, Audrey Low, Weilan Wang, B.T. Thomas Yeo, Yanhong Dong, Nagaendran Kandiah, Geert J. Biessels, Andrea B. Maier

## Abstract

Hallmarks of biological ageing are closely interconnected with neurodegenerative diseases. Deoxyribonucleic acid methylation (DNAm) clocks quantify biological age and may serve as peripheral biomarkers of neurodegenerative diseases. This systematic review and meta-analysis aim to examine associations between DNAm-based biological age metrics and neurodegenerative diseases. Six databases were systematically searched from 1^st^ January 2011 to 15^th^ April 2026 for studies reporting associations between DNAm-based ages and neurodegenerative diseases including all-cause dementia, Parkinson’s Disease, Multiple Sclerosis, and Amyotrophic Lateral Sclerosis. Correlated-and-hierarchical-effects meta-analyses were fitted using restricted maximum likelihood with Knapp-Hartung adjustment. Of 42,023 records screened, 37 observational studies (n=46 365; 60.5% female; median of mean age 69.4 years [IQR 62.5-73.3)) and seven Mendelian randomization studies met inclusion criteria. In longitudinal meta-analyses, higher GrimAge epigenetic age acceleration (EAA) was significantly associated with higher dementia risk (HR per SD 1.11 [95% CI 1.02–1.20]; I² = 0.0%; p = 0.018), whereas Horvath1 EAA, Hannum EAA, PhenoAge EAA, and DunedinPACE were not significantly associated with dementia risk. Evidence for other neurodegenerative diseases were heterogeneous across clock generations and study designs. Mendelian randomization provided limited evidence for causality. Higher GrimAge EAA was the only DNAm age measure consistently predictive of higher all-cause dementia risk. Overall, however, DNAm age clocks show limited and inconsistent associations with neurodegenerative diseases, providing insufficient support for their use as disease biomarkers. These findings highlight the need to develop and independently validate disease-specific DNAm clocks and undertake pathway analyses to identify methylation changes associated with neurodegeneration and the biological processes they reflect.

## Introduction

The prevalence of neurodegenerative diseases are expected to rise with global demographic changes and increasing life expectancy, imposing escalating medical and economic burden on public healthcare^1^. Major neurodegenerative diseases, including Alzheimer’s disease (AD), Parkinson’s disease (PD), Multiple Sclerosis (MS), and Amyotrophic lateral sclerosis (ALS) contribute to cognitive, motor and behavioural impairments, and are leading causes of reduced disability-adjusted life years, institutionalization, and mortality worldwide^2, 3^. Among the multiple risk factors for neurodegeneration, the ageing process itself has by far the most impact, with cellular and molecular hallmarks of biological ageing being closely linked to neurodegenerative diseases and their clinical course^4, 5^. Given individual variability in ageing trajectories, biomarkers capable of quantifying the ageing process or determining biological age may provide novel opportunities in mechanistic insights, risk stratification, diagnosis and prognostication of neurodegenerative diseases^6, 7^.

Among the most established biological ageing clocks are deoxyribonucleic acid methylation-based (DNAm) clocks, which utilize machine learning algorithms trained with DNAm patterns at cytosine-phosphate-guanine sites within the genome to quantify DNAm age of tissues or cells^8, 9^. First-generation DNAm clocks were calibrated based on chronological age as the outcome while second-generation clocks were trained on clinical parameters linked to morbidity and mortality^10^. Third-generation clocks quantified pace of ageing across various organ systems and newer ‘explainable’ clocks emphasized biological interpretability^11–13^. The difference between DNAm age and chronological age is referred to as epigenetic age acceleration (EAA) or age deviation. Age deviation is calculated as the difference between DNAm age and chronological age while EAA is derived as the residual from regressing DNAm age on chronological age. The application of DNAm clocks trained on peripheral biosamples is compelling in providing a minimally invasive approach for longitudinal monitoring of neurodegenerative processes.^14^ However, its clinical utility for risk stratification and prediction of neurodegenerative diseases is unclear as existing evidence remains fragmented across different DNAm clocks, neurodegenerative diseases and study designs^15^. Although previous reviews have summarized associations between DNAm clocks and neurodegenerative diseases, the rapidly expanding diversity of DNAm measures have not been quantitatively synthesized^16–18^. Consequently, important uncertainties remain whether DNAm EAA is associated with established neurodegenerative disease, whether they predict incident diseases, and whether the observed associations may reflect causal processes.

Accordingly, the primary aim of this systematic review and meta-analysis was to provide a comprehensive evaluation of the cross-sectional and longitudinal associations between DNAm clocks and neurodegenerative diseases.^17, 18^ Its secondary aim was to synthesize Mendelian randomization studies to examine whether genetically proxied DNAm age may have a causal role in neurodegenerative disease risk.

## METHODS

The review is part of the Global Epigenetic Age Consortium^19^, an international collaboration dedicated to synthesizing evidence on the association of DNAm clocks with clinically relevant outcomes including frailty^20^, physical activity^21, 22^, reproductive history^23^, and clonal hematopoiesis^24^, with aims of advancing the potential clinical translation of DNAm clocks for geroscience and precision geromedicine^25^.

### Data Sources and Search strategy

This systematic review and meta-analysis followed the Preferred Reporting Items for Systematic reviews and Meta-Analyses (PRISMA) guideline^26^. The protocol was registered with the International Prospective Register of Systematic Reviews (PROSPERO; CRD42024499021). Searches included articles published between 1^st^ January 2011 and 15^th^ April 2026 as the first DNAm clock had been published in 2011^27^. The search was performed in six electronic databases, including Embase, Cochrane Central Register of Controlled Trials (CENTRAL), PubMed, Ovid, Scopus and Web of Science, using MeSH terms and keywords related to DNAm clocks (Supplementary Table 1). The search was constructed with the assistance of a senior librarian from the National University of Singapore. Additional studies were searched using the reference list of retrieved articles. Authors were contacted for additional data to verify eligibility for inclusion.

### Article selection

Two assessors (JT and WW) independently removed duplicates and screened the titles and abstracts of articles for eligibility using the Covidence (Veritas Health Innovation, Melbourne, VIC, Australia). Full-text articles were screened independently (by JT and WW) and organised using the tagging feature of Covidence into specific age-related diseases, conditions and health outcomes. Epigenetic clocks not based on DNAm such as assay for transposase-accessible chromatin (ATAC) clock^28^ and the histone clock^29^ were excluded. Disagreements were resolved by a third assessor (ABM). The inclusion criteria consisted of peer-reviewed population-based cohort or case-control studies written in English involving adults (mean/median age ≥ 18 years) from general populations or disease-specific populations. Studies were eligible if they (1) assessed cross-sectional or longitudinal associations between DNA methylation age and neurodegenerative diseases, (2) included DNAm clocks expressed as DNAm age, DNAm age deviation (AgeDev, the difference between DNAm age and chronological age), or epigenetic age acceleration (EAA; derived using residuals from a linear model regressing DNAm age on chronological age in control subjects) (3) measured DNAm age using minimally invasive sample collection methods, including blood, saliva, buccal cells, or skin, (4) included neurodegenerative diseases such as dementia or any subtype of dementia (i.e. Alzheimer’s disease (AD), Lewy body dementia (LBD), and frontotemporal dementia (FTD))^30, 31^, PD, MS, and ALS, that are assessed based on established clinical criteria including the International Classification of Diseases 11^th^ Revision (ICD-11 – Chapter 08 Diseases of the nervous system)^32^ and Diagnostic and Statistical Manual of Mental Disorders, fifth edition, text revision (DSM-5-TR)^33^ (eMethods). MR studies that evaluated associations of DNAm clocks and neurodegenerative diseases were also included.

### Data Extraction

Two assessors (ZK and JT) independently extracted data, including title, first author, year of publication, study design, cohort location, and participants’ characteristics (sample size, group size, age, and sex), DNA source, DNAm profiling method (including whole genome bisulfite sequencing, Illumina array or other technology), DNAm clock employed, effect estimates and the adjustment models applied. For MR studies, information on the MR design (one-sample or two-sample; unidirectional or bidirectional), inverse-variance weighted (IVW) estimates, sensitivity analyses conducted such as MR-Egger regression, weighted median and weighted mode, statistical significance (p-value) and multiple testing correction (Bonferroni or Benjamini-Hochberg false discovery rate) were extracted. Nominal significance (p < .05) from the IVW estimates were used for synthesis in the systematic review. Findings that do not survive the primary study’s own correction for multiple comparisons or was inconsistent across MR sensitivity methods within the same study, were explicitly reported.

### Risk-of-Bias Assessment

Risk of bias was assessed by two independent raters (ZK and AL) using the Newcastle-Ottawa Quality Assessment Scale^34^ for case-control and cohort studies. Quality of MR studies were evaluated using the Strengthening the Reporting of Observational Studies in Epidemiology using Mendelian Randomization (STROBE-MR) statement^35^.

### Meta-analysis

#### Study Eligibility and Effect-size Harmonization

Meta-analysis was restricted to studies reporting a hazard ratio (HR) with a 95% CI for incident all-cause dementia and was conducted separately for each DNAm clock. As studies reported effects on different scales, all estimates were harmonised to the HR per 1 SD higher EAA. Studies publishing no usable SD, and whose effect could therefore not be placed on a common scale, were excluded.

#### Statistical model

Several cohorts contributed more than one estimate to the same pooled analysis. Such estimates are not independent. The correlated-and-hierarchical-effects (CHE) working model was therefore used^36^, fitted with metafor^37^. This combines a three-level random-effects structure, allowing true effects to vary both between and within studies, with a block-diagonal sampling variance-covariance matrix assuming a constant correlation of rho=0·6 among the sampling errors of estimates from the same cohort. A three-level model alone does not address the latter, because the sampling variance-covariance matrix is treated as diagonal unless its off-diagonal elements are supplied. Models were fitted by restricted maximum likelihood with the Knapp-Hartung adjustment, and pooled log HRs were back-transformed for presentation. Cluster-robust (CR2) SEs with Satterthwaite degrees of freedom were computed alongside every model. Where no cohort contributed more than one estimate to a clock family, the within-study variance component is not identifiable, and a two-level model was fitted instead. All hypothesis tests were two-tailed, with statistical significance set at alpha=0·05; the comparison of interest in every model was the pooled per-SD hazard ratio against the null of HR=1 for each DNA methylation clock.

#### Heterogeneity, robustness and small-study effects

Between-study heterogeneity was quantified with Cochran’s Q, tau-squared, and a multilevel I^2^ partitioned into between-study and within-study components^38^, with I^2^ flagged where fewer than five estimates contributed, since at small k it is dominated by k itself^39^. 95% prediction intervals are reported alongside each pooled estimate to indicate the range in which the effect in a new cohort would be expected to fall^40^. Robustness was examined by varying the assumed within-study sampling correlation (rho=0·3 and 0·9), by clustering on cohort rather than on publication, by omitting the smaller of the two overlapping WHIMS reports, by reinstating the excluded odds-ratio study, and by leave-one-study-out analysis. Influence was assessed with Baujat plots, DFFITS, Cook’s distance, and leave-one-out heterogeneity. Small-study effects were assessed with contour-enhanced funnel plots, Egger’s regression restricted to clock families with at least ten estimates^41^, and trim-and-fill applied to a one-estimate-per-study comparator; trim-and-fill imputes hypothetical studies and is reported as a sensitivity analysis only. Analyses were done in R (version 4·6·1) with metafor (version 4·8·0) and clubSandwich.

#### Covariates

Associations were analysed at two levels of covariate adjustment. The minimally adjusted analysis used each study’s least-adjusted model, which controlled for at least chronological age and sex. The fully adjusted analysis used each study’s most fully adjusted model.

## RESULTS

### Study characteristics

The literature search identified 40 023 records, of which 24 896 duplicates were removed, and a total of 15 437 records were excluded based on titles and abstracts. Of the 1690 full-text articles assessed, 1644 were excluded, leaving 44 studies, of which eight cohorts were included in the meta-analysis (Figure 1). Overall, 37 observational studies and seven MR studies were included. Dementia was examined in 29 studies^12, 13, 42–67^, PD in 13 studies^13, 45, 47, 50, 50, 60, 64, 68–74^, MS in eight studies^45, 47, 64, 75–79^, and ALS in five studies^47, 60, 80–82^ (Table 1).

**Figure 1.**
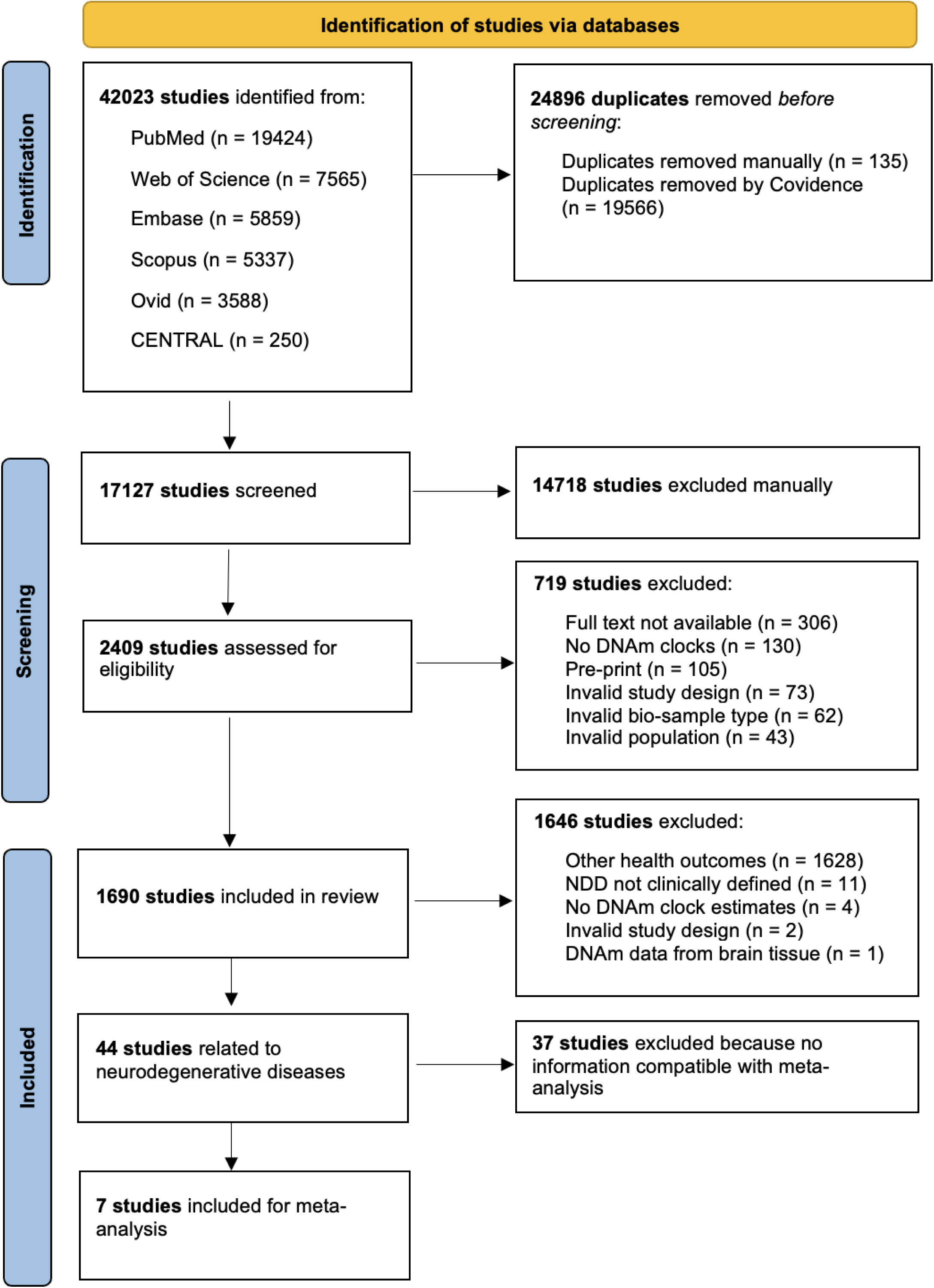
Flow diagram of search strategy. Note: CENTRAL = Cochrane Central Register of Controlled Trials.

**Table 1.**
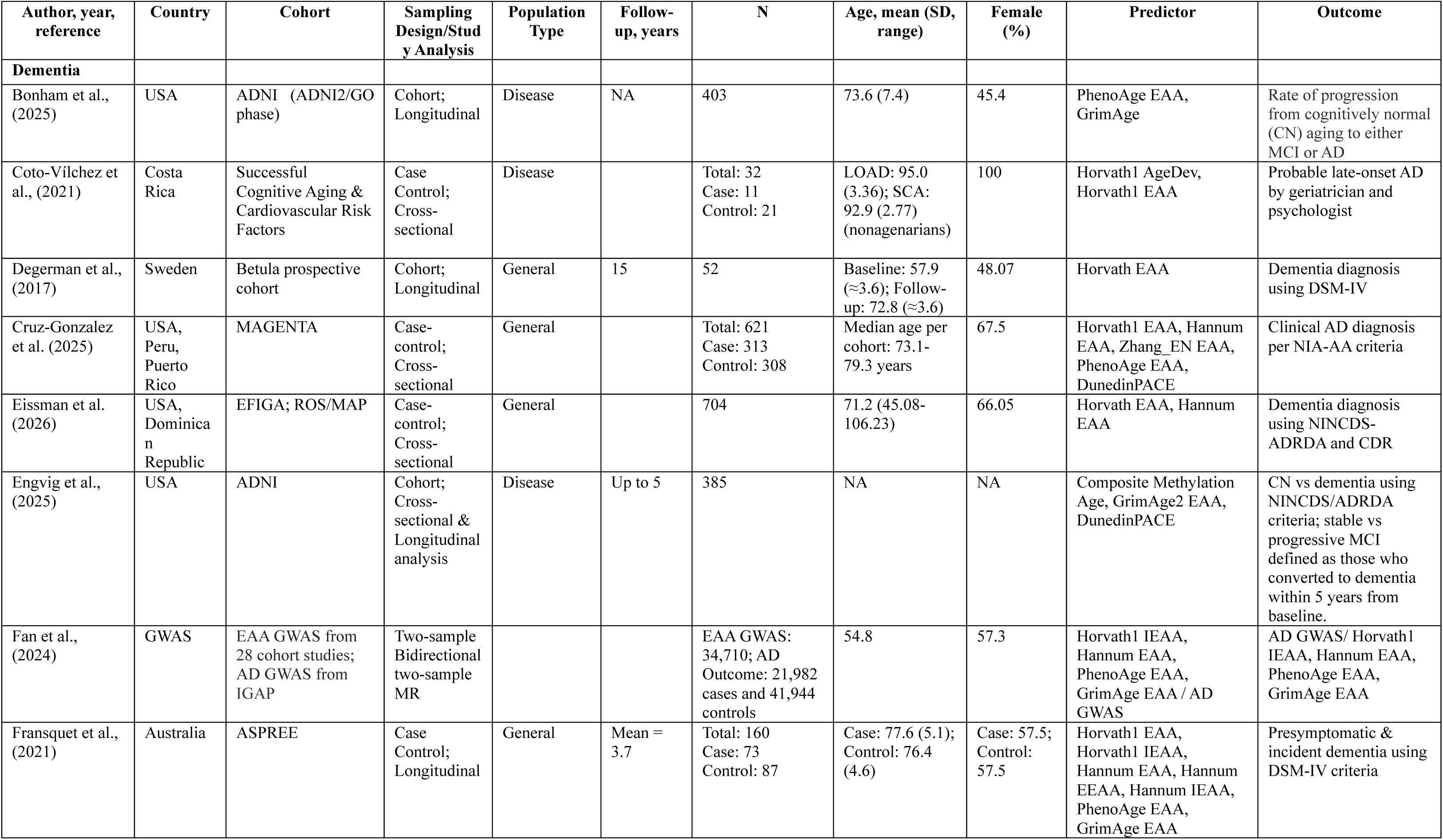
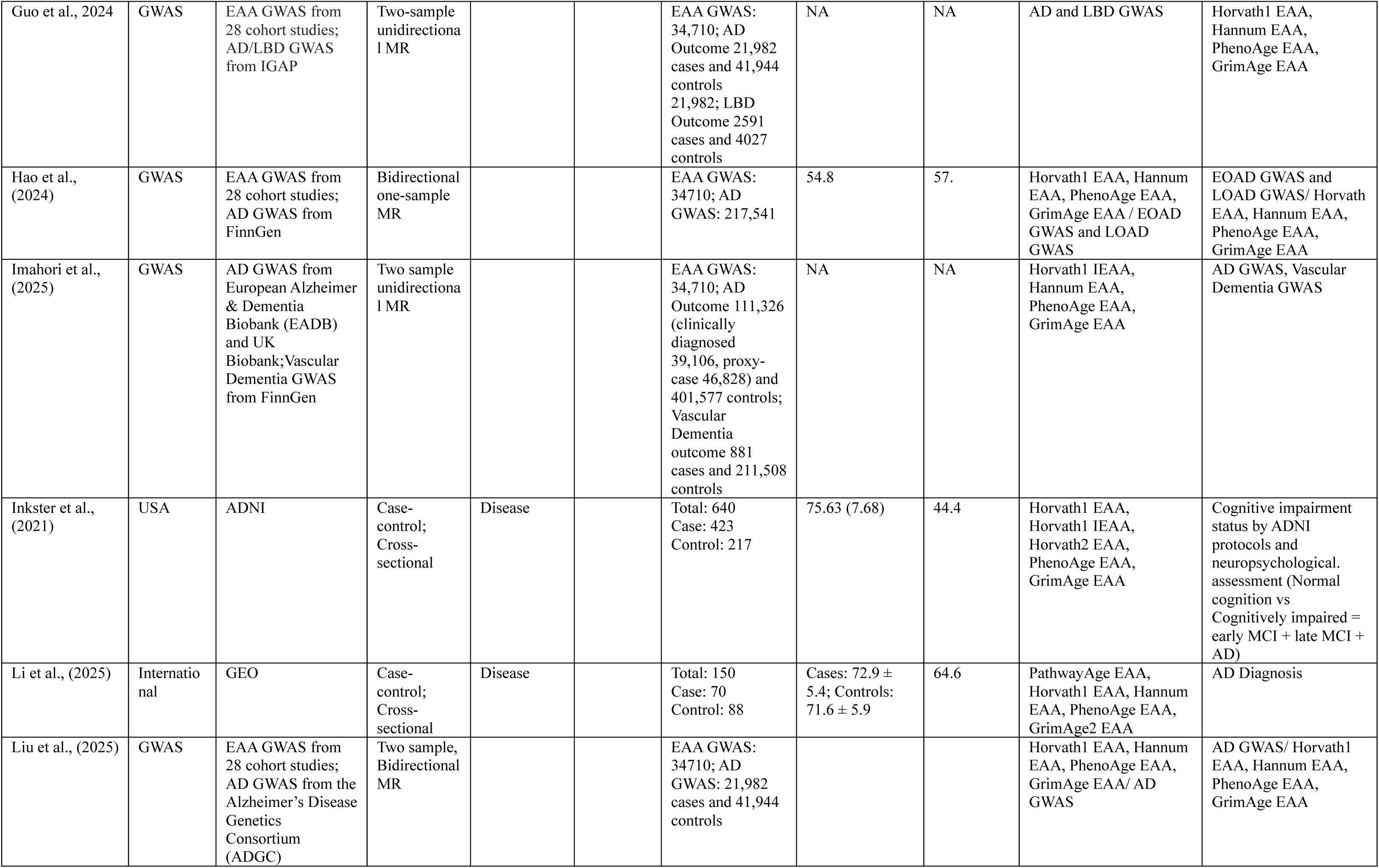
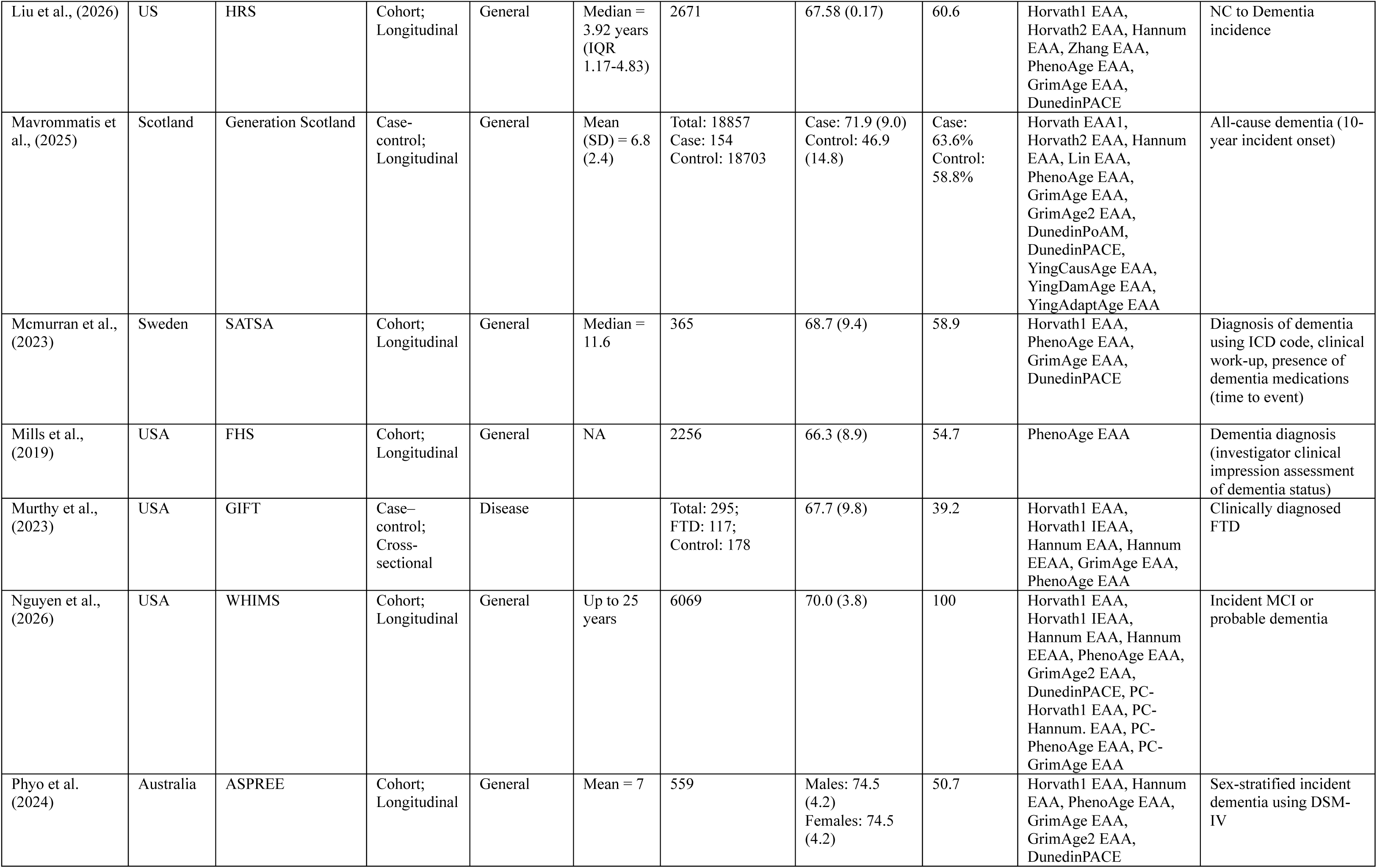
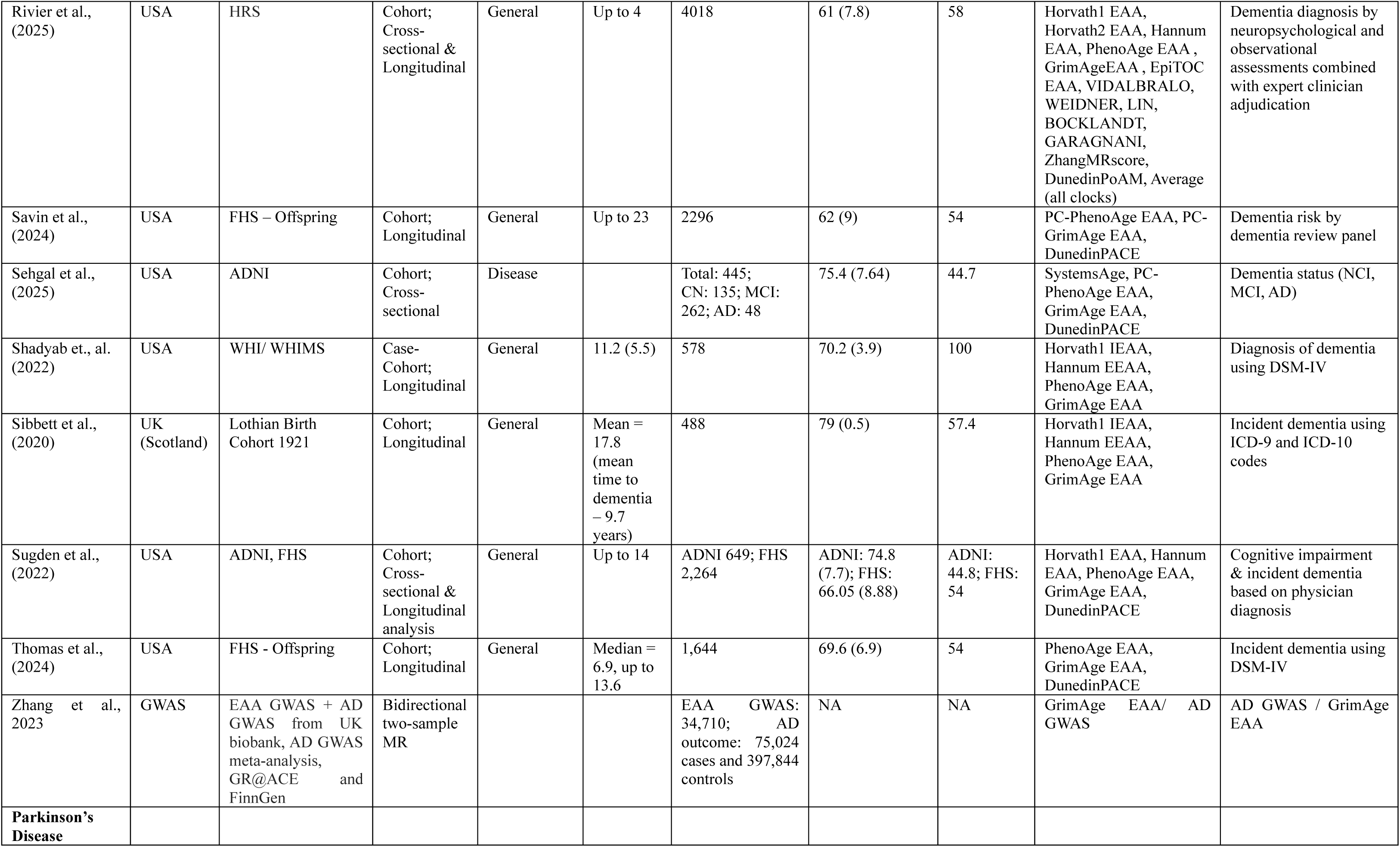
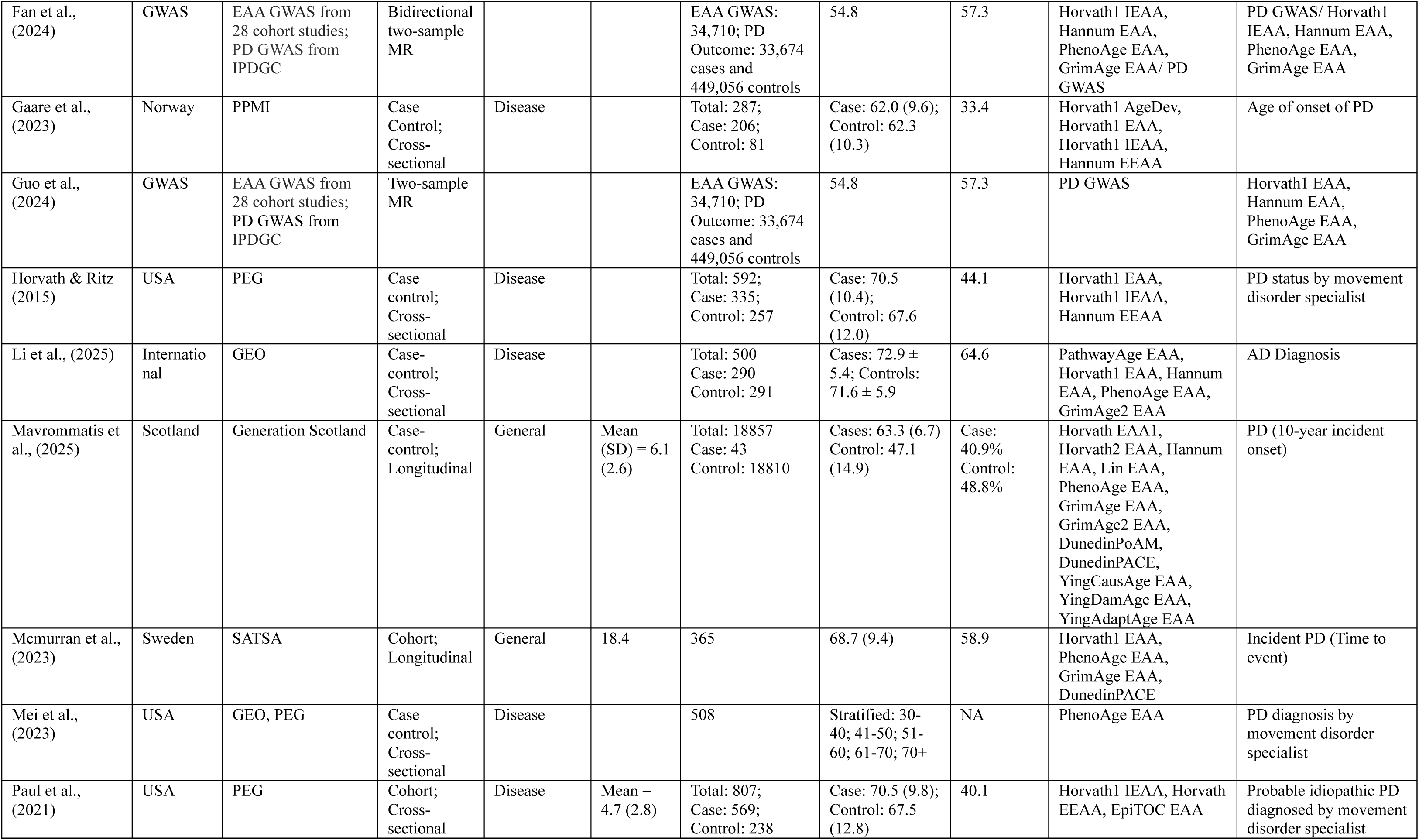
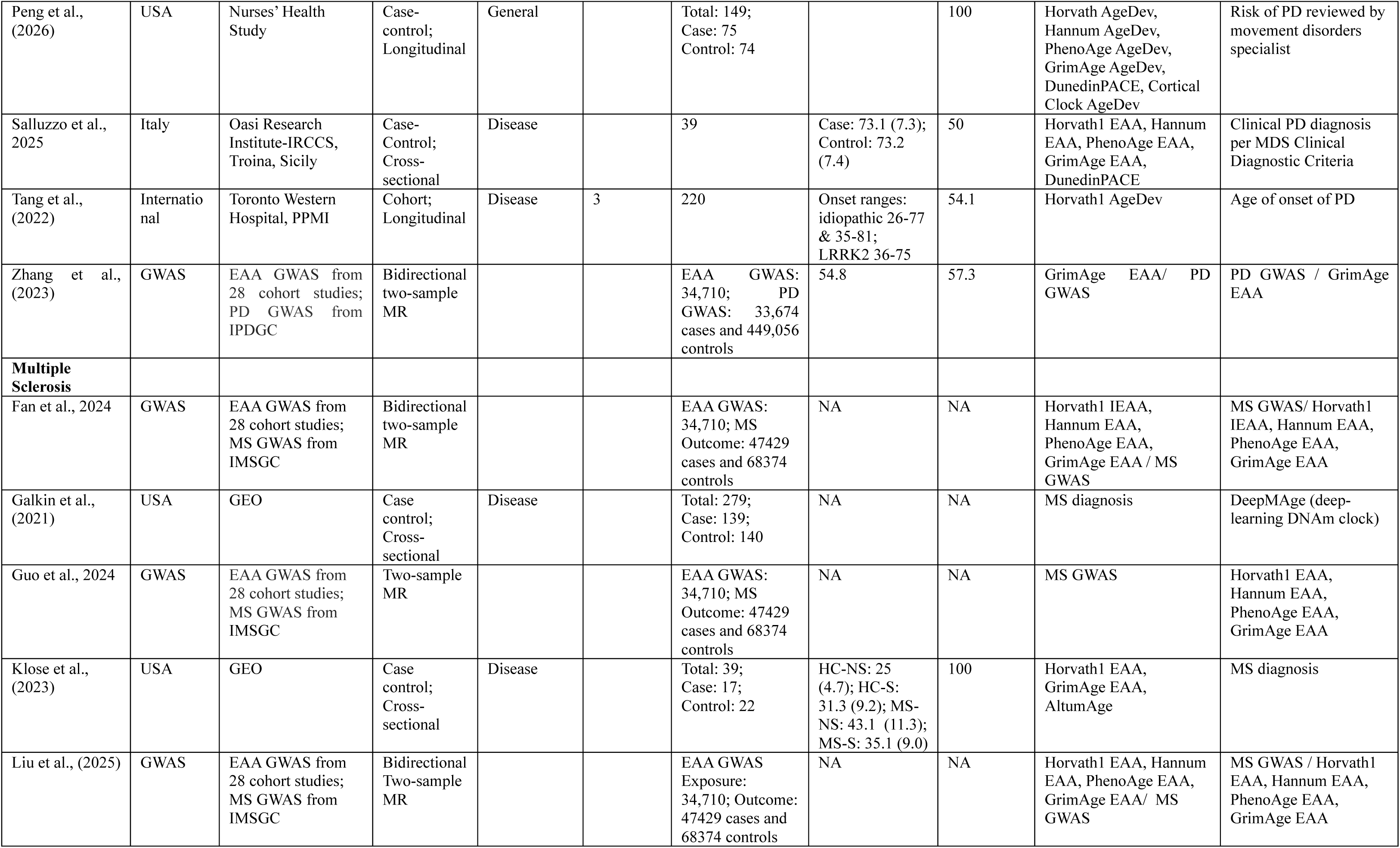
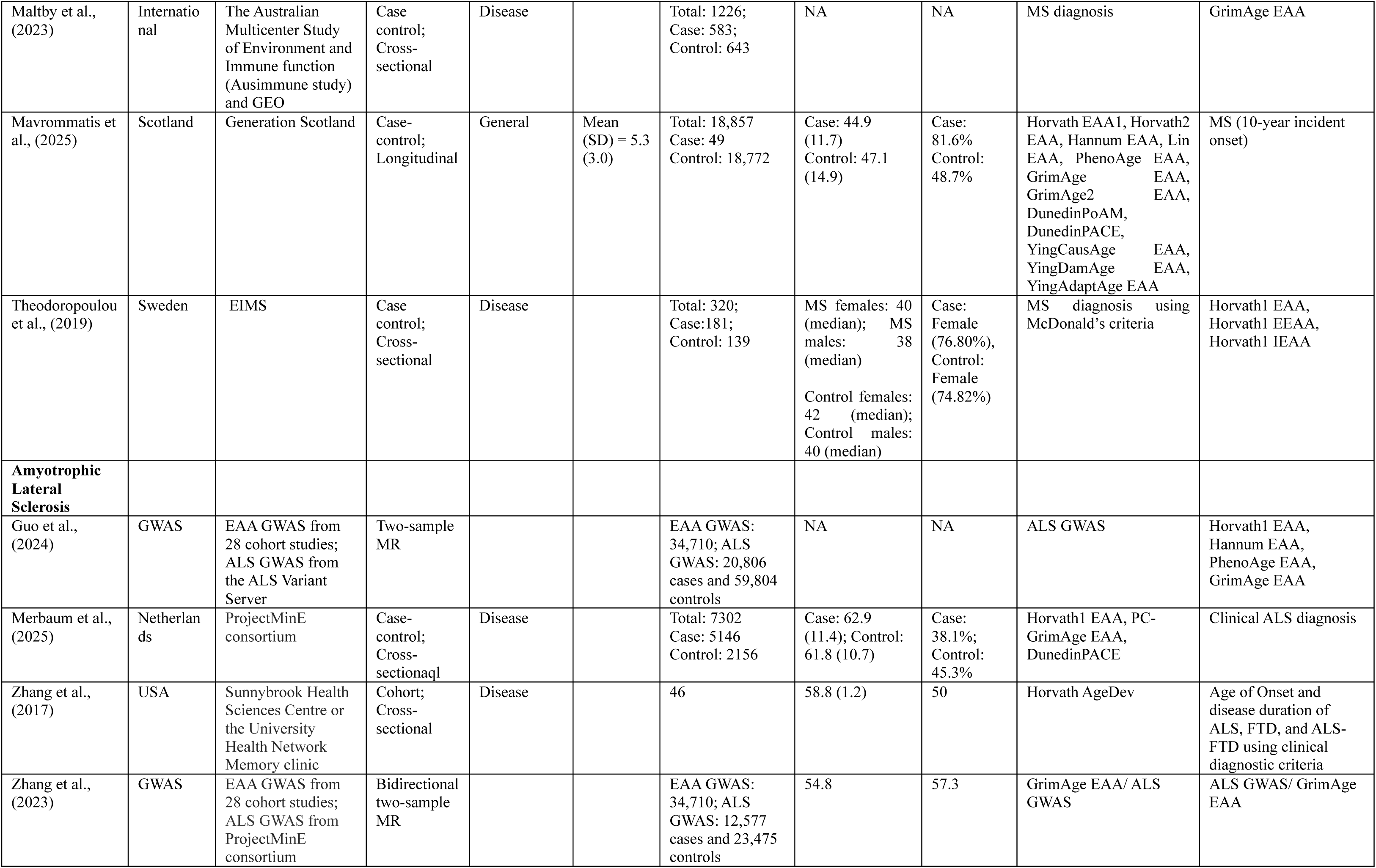
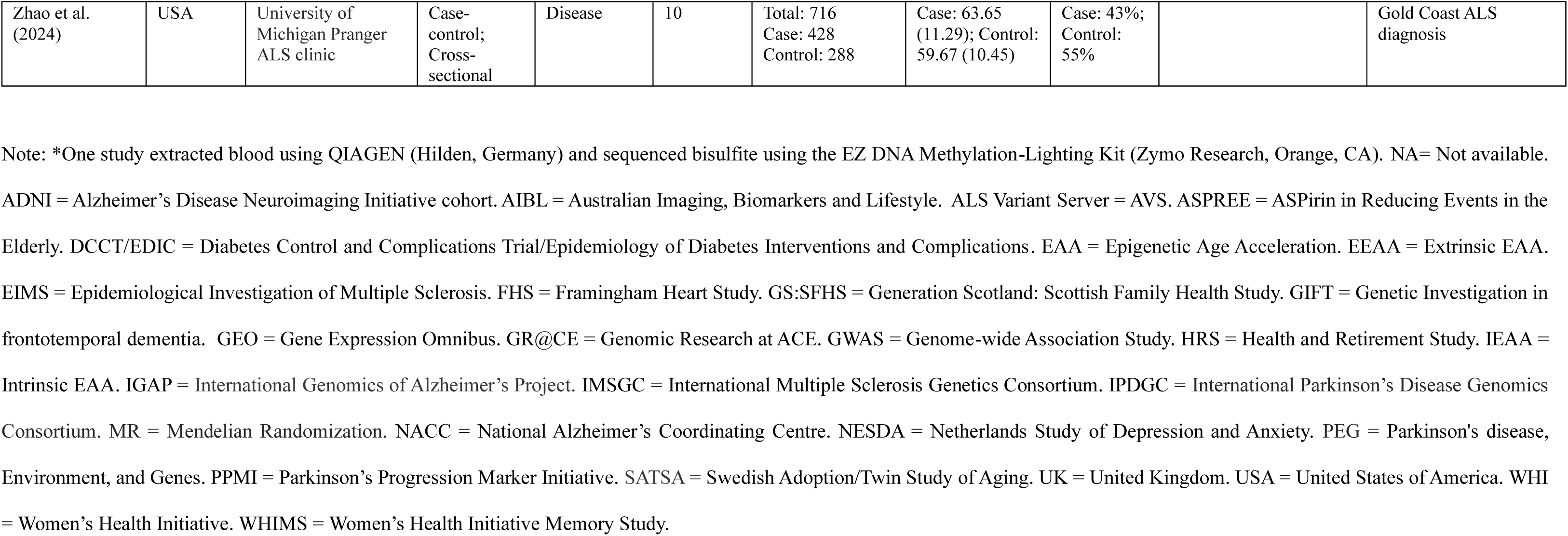
Characteristics of included studies in the systematic review.

Of the 37 observational studies included, 21 were based in North America, followed by Europe (n=8), Oceania (n=2), and Latin America (n=1), while the remaining five used multi-site or international datasets (Figure 2). The following cohorts were frequently reported more than once: Alzheimer’s Disease Neuroimaging Initiative (ADNI, n=5), Gene Expression Omnibus (GEO=5), Framingham Heart Study (FHS, n=4), Parkinson’s disease, Environment, and Genes (PEG, n=3), Health and Retirement Study (HRS, n=2), Women’s Health Initiative Memory Study (WHI/WHIMS, n=2), ASPirin in Reducing Events in the Elderly study (ASPREE, n=2), and Parkinson’s Progression Markers Initiative (PPMI, n=2). After deduplicating participants shared across overlapping cohorts, the total sample comprised 46 365 unique individuals across cohorts (60.5% females, median of mean age=69.4 [IQR 62.5-73.3])), with individual sample sizes ranging from 32 to 18 857 individuals. Among the observational studies, 18 were cohort studies and 19 were case-control studies, with 19 studies employing cross-sectional analysis (45.2% females, median of mean age=69.6 [IQR 62.6-73.2])), 15 involving longitudinal analyses (65.4% female, median of mean age=69.6 [IQR=66.3-73.6]), and three using both approaches.

**Figure 2.**
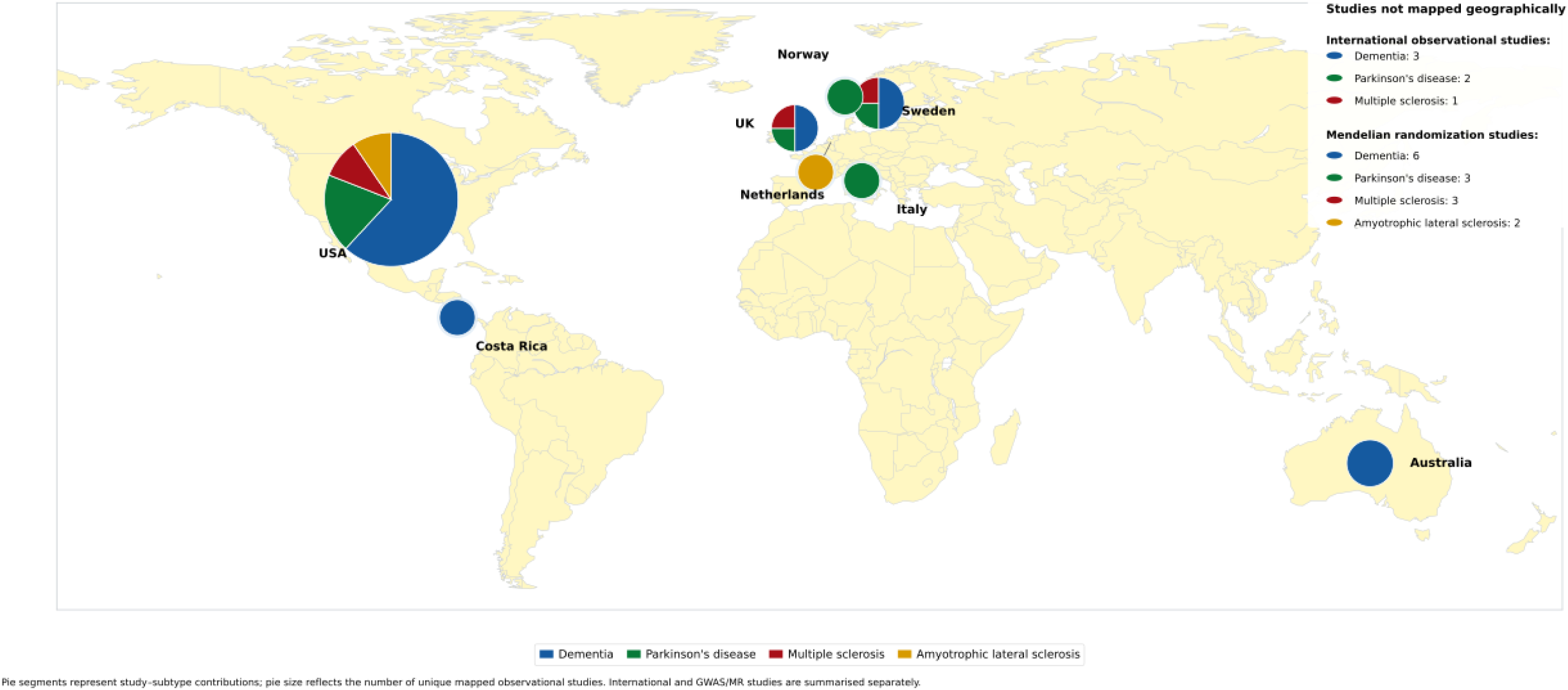
World map plot of studies examining deoxyribonucleic acid methylation-based clocks in neurodegenerative diseases Note: World map plot, displaying data sources distribution by country of origin and subtype of neurodegenerative disease. Bubble size reflects number of observational studies per country. Multinational observational and Mendelian Randomization studies are shown in the inset table. Countries filled with light-yellow colour are not featured in any of the reviewed publications. This map plot is based on the data from Table 1.

Seven MR studies leveraged Genome-Wide Association Study (GWAS) summary statistics from cohort and biobank databases to test putative causal relationships between DNAm age and neurodegenerative diseases^45, 47, 48, 60, 63, 66, 79^. All seven used a two-sample design, of which four were bidirectional and three unidirectional. These analyses involved predominantly European-ancestry summary datasets (e.g., AD: up to 111 326 cases and 401 577 controls; vascular dementia: up to 888 cases and 211 508 controls; PD: 33,674 cases and 449 056 controls; MS: 47 429 cases and 68 374 controls; ALS: 20 806 cases and 59 804 controls) that were assessed with an EAA GWAS cohort of 34 710 individuals (57.3% females, mean age=54.8 years)^83^.

First-generation DNAm clocks included Horvath1^84^ (n=30)^13, 43, 44, 46, 48–50, 52–54, 56–58, 62–66, 68, 69, 72– 74, 76, 78–80, 82, 85, 86^, Horvath2 87 _(n=5)_49, 54, 64, 76, 86_, Hannum_88 _(n=23)_13, 45, 46, 48, 52–54, 56–58, 62–66, 68, 69, 74, ^76, 78, 79, 85, 86^ and Zhang EN/BLUP^89^ (n=2)^13, 85^. Second-generation clocks comprised of _PhenoAge_90 _(n=28)_13, 42, 45, 46, 48–59, 63–66, 70, 73, 74, 76, 78, 79, 85, 86_, GrimAge_91 _(n=26)_12, 42, 45, 46, 48–50, 52–60, 63, 64, 66, 73, 74, 76, 77, 79, 81, 86_, GrimAge2_92 _(n=5)_13, 53, 64, 65, 93_, and ZhangMRscore_89_(n=2)_54, 64_, and_ EpiTOC (mitotic age, n=2).^54, 71^ Third-generation clocks consisted of DunedinPACE^94^ (n=15)^12, 50, 53, 55, 58, 59, 64, 65, 73, 74, 82, 85, 86, 93^ and DunedinPoAm^11^ (n=2)^54, 64^, while newer clocks included SystemsAge^12^ and PathwayAge. Principle-component clocks were investigated in four studies^12, 82, 85, 95^. Newer clocks that were examined included SystemsAge (n=1)^12^, Pathway Age (n=1)^13^, DamAge^96^ (n=2)^13, 64^, AdaptAge^96^ (n=2)^13, 64^, Cortical clock (n=1), AltumAge^76^ (n=2)^76^,

DeepMAge^75^ (n=1)^75^, a derivative clock of composite methylation age^67^ (n=1)^67^ and ensemble clocks (n=2)^54, 85^. Cell composition effects were addressed in 24 studies through cell-independent _measures_46, 49, 52, 56, 57, 68, 69, 78, 85, 97_, covariate adjustments_13, 43, 44, 53, 55, 58, 59, 64, 65, 71, 72, 76, 82_, or cell-_specific analyses.^77^ Detailed study characteristics and effect sizes are given in Supplementary Tables 2-5. Of the 18 cohort studies, 16 were rated as high quality^12, 42, 44, 50, 53–59, 65, 71, 72, 86, 93^ while two were rated as moderate quality^51, 80^ (Supplementary Table 6). Of the 19 case-control studies evaluated, 15 were rated as high quality^13, 46, 49, 52, 62, 64, 69, 70, 73, 76–78, 81, 82, 85^ and four were rated as moderate quality^43, 70, 74, 75^. All seven MR studies were rated as high quality (Supplementary Table 7).

### Dementia

Twenty-three observational studies included general populations (n=42 277 individuals) and dementia populations (n=4323), and four MR studies using the same EAA GWAS with different AD GWAS (sample sizes ranging from 63 926 to 472 868 individuals).

In cross-sectional analyses of general populations (n=2), dementia diagnosis was significantly associated with higher EpiTOC EAA, GrimAge EAA and ZhangMRScore EAA^54^ whereas there were no associations with the EAA of other DNAm age including Horvath1, Horvath2, Hannum, Vidalbralo, Weidner, Bocklandt, Garagnani, Lin, PhenoAge, ZhangMRscore, or DunedinPoAM^46, 54^ (eFigure 1).

In cross-sectional analyses of disease populations (n=9), positive associations with AD diagnosis were reported for composite methylation age^93^, PathwayAge^13^, AdaptAge^13^, and SystemsAge^12^ (eFigure 2). Mixed evidence was observed for the associations between Hannum EAA (3^13, 58, 85^/5 studies)^52, 62^, PhenoAge EAA (3^49, 52, 58^/5 studies)^13, 85^, DunedinPACE (2^58, 85^/4 studies)^12, 13^ and AD diagnosis, whereas there were no associations between Horvath1 Agedev^43^, Horvath1 IEAA^49^, Horvath1 EAA (5^13, 43, 49, 52, 58^/7 studies)^62, 85^, Horvath2 EAA^49^, PC-PhenoAge EAA^12^, GrimAge EAA (5/5 studies)^12, 13, 49, 52, 58^, YingDamAge^13^, or other Systems scores^12^ with the clinical diagnosis of AD or FTD.

In longitudinal analyses across general populations (n=13), Horvath1 AgeDev^44^, GrimAge EAA (6^50, 53, 55, 57–59/10^ studies)^46, 54, 56^, GrimAge2 EAA (3/3 studies)^53, 64, 65^, PC-GrimAge EAA^65^, were positively associated with all-cause dementia risk (Figure 3; eFigure 3). Mixed evidence was observed for the associations between PhenoAge EAA (5^53, 54, 56, 57, 65, 86^/11 studies) ^51, 58, 59, 64, 98^, PC-PhenoAge EAA (1^55^/2 studies^65^), DunedinPoAM (1^64^/2 studies)^54^, DunedinPACE (4^55, 58, 59, 65^/8 studies)^53, 64, 86, 98^ and dementia risk. No associations in longitudinal analyses were observed for Horvath1 EAA (73^13, 53, 54, 56, 57, 64, 65^/9 studies)^58, 98^, Horvath1 IEAA (3/3 studies)^56, 57, 65^, Horvath2 (2^54, 64^/2 studies)^86^, Hannum EAA (5^53, 58, 64, 65, 86^/6 studies)^54^, Hannum EEAA (3/3 studies)^56, 57, 65^, EpiTOC (2/2 studies)^54, 64^, Vidalbralo^54^, Weidner^54^, Bocklandt^54^, Garagnani^54^, Lin^54^, ZhangMRscore^54^, AdaptAge^64^, and DamAge^64^ with all-cause dementia risk. In a population-based cohort following 400 participants with mild cognitive impairment at baseline over four years, higher Horvath2 EAA was significantly associated with increased risk of conversion to all-cause dementia (33.0% conversion over a median 3.58-year follow-up; HR = 1.62, 95% CI 1.09-2.40), whereas other DNAm clocks were not associated with risk of MCI-to-dementia conversion^86^.

**Figure 3.**
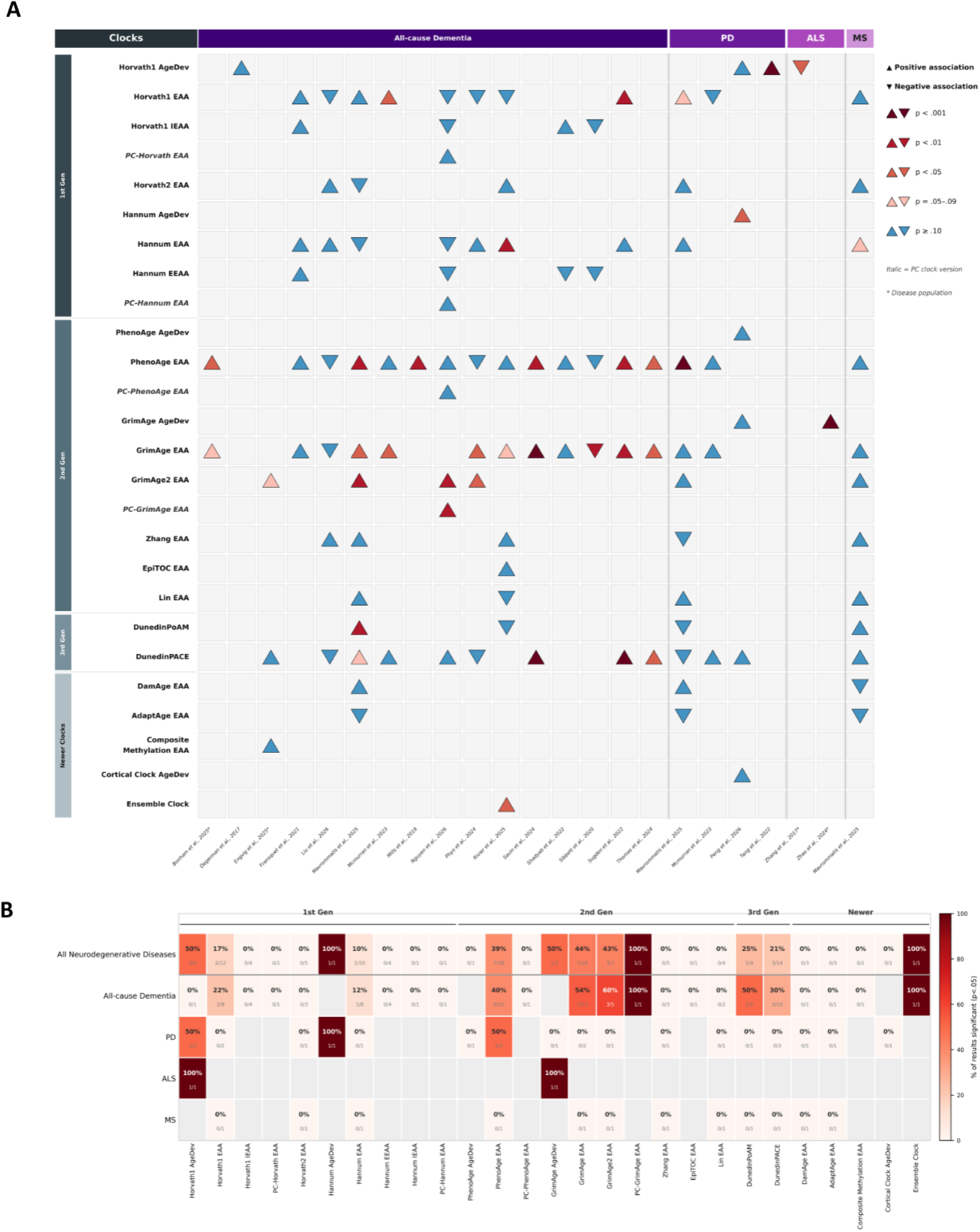
Evidence map of DNAm clock associations with neurodegenerative diseases across longitudinal studies. Note: Each cell represents the association between an epigenetic clock (y-axis) and neurodegeneration outcome within a given study (x-axis), where data were available. Triangle orientation indicates the direction of association (▴ positive; ▾ negative), and colour intensity reflects the level of statistical significance: dark crimson (p < .001), dark red (p < .01), medium red (p < .05), pale pink (p = .05–.09), and blue (p ≥ .10). Where multiple estimates were reported within a study (e.g., sex-stratified analyses), the estimate with the smallest p-value was displayed. Studies are grouped by disease domain (All-cause Dementia, Parkinson’s Disease (PD), Amyotrophic Lateral Sclerosis (ALS), and Multiple Sclerosis (MS)) and clocks are grouped by generation (1^st^ generation, 2^nd^ generation, 3^rd^ generation and newer clocks). *Disease cohort studies with pre-existing diagnosis of neurodegenerative diseases at baseline.

In longitudinal analyses of a disease population, PhenoAge EAA was positively associated with risk of AD while GrimAge EAA showed no association (eFigure 4).^42^ There were no significant longitudinal associations between GrimAge2, composite methylation age or DunedinPACE with the conversion from MCI to AD^93, 99^.

**Figure 4.**
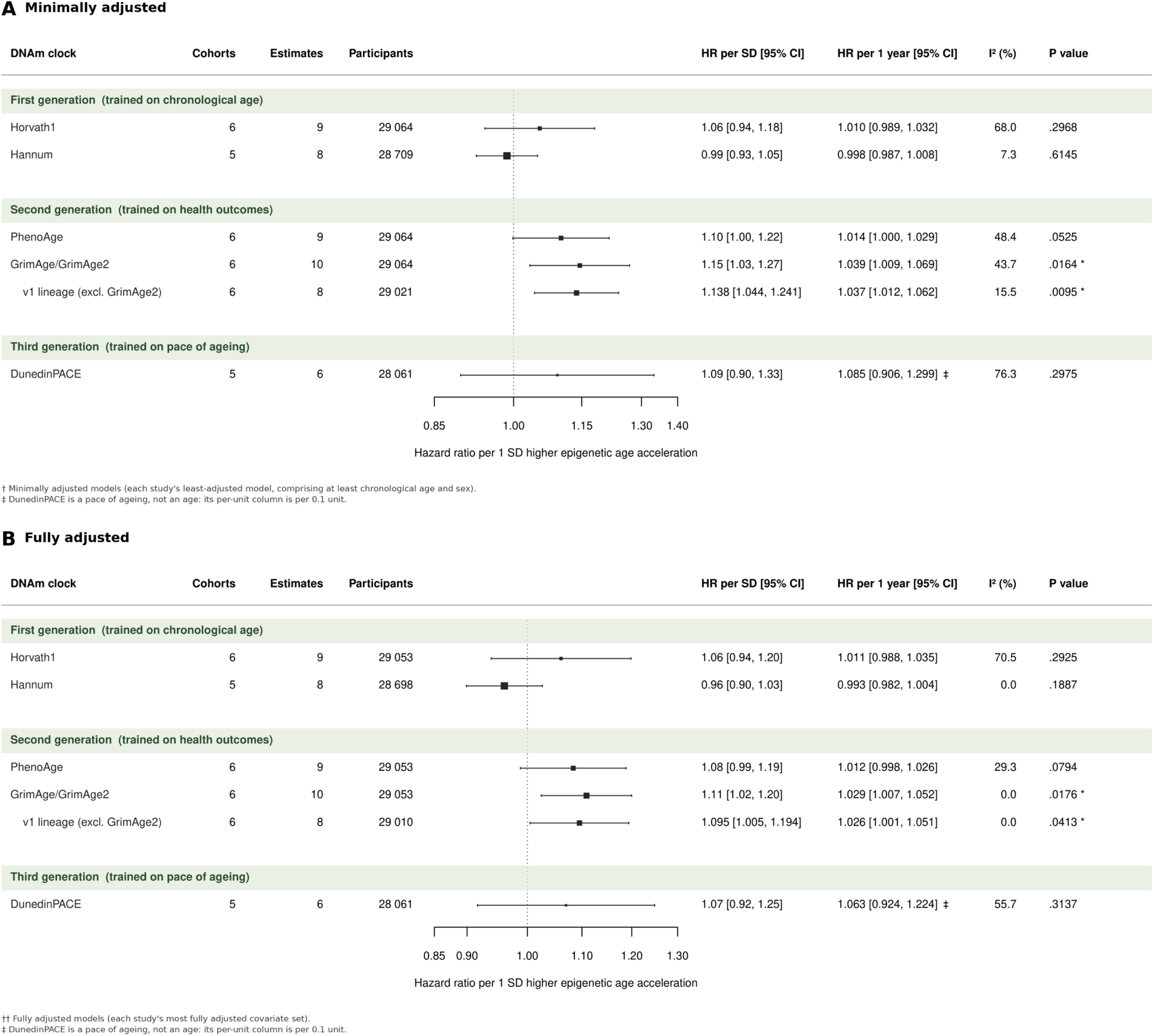
Meta-analytic association between GrimAge EAA and all-cause dementia. (A) Fully-adjusted models between GrimAge EAA (inclusive of GrimAge EAA, GrimAge2 EAA, PC-GrimAge EAA) and all-cause dementia. (B) Basic-adjusted models between GrimAge EAA and all-cause dementia. Cohorts refer to the number of independent cohorts contributing to each pooled estimate. Estimates is the number of effect estimates pooled.

Across MR studies, genetically predicted EAA was not associated with AD or vascular dementia risk for Horvath1 IEAA (2/2 studies)^45, 66^, Horvath EAA (2/2 studies)^48, 63^, Hannum EAA (3^45, 63, 66^/4 studies), PhenoAge EAA (4/4 studies)^45, 48, 63, 66^ and GrimAge EAA (4^45, 48, 60, 66^/5 studies). Two exceptions were noted. Hannum EAA was causally associated with increased risk of late-onset AD, but not early-onset AD^48^. GrimAge was causally associated with increased risk of AD by IVW (OR=1.025, 95% CI 1.006-1.004, p=.009), although this estimate was not robust to MR-Egger sensitivity analysis (OR=1.021, 95% CI 0.946-1.102, p=0.618), potentially owing to its small and underpowered instrument count (7 SNPs for GrimAge, 15.9% power)^63^. In the reverse direction, negative causal associations between genetically predicted AD and GrimAge EAA was replicated across two studies, although neither survived within-study Bonferroni corrections. AD had no causal effects on other DNAm clocks^45, 47, 48^.

### Parkinson’s Disease

For PD, ten observational studies included general population cohorts (n=19 222 individuals) and PD populations (n=3102), and three MR studies using identical PD GWAS dataset (n=482 730). In cross-sectional analyses within PD populations, PD diagnostic status was significantly associated with higher Hannum EEAA (2^69, 71^/3 studies)^68^, EpiTOC EAA^71^, DunedinPACE^13^, and PathwayAge (Top 20 disease pathways)^13^. On the other hand, Horvath IEAA (2^68, 69^/3 studies)^71^, Hannum EAA^13^, PhenoAge AgeDev^70^, PhenoAge EAA^13^, GrimAge2 EAA^13^, DamAge EAA^13^, and AdaptAge EAA^13^ showed no differences between PD patients and controls. Horvath1 AgeDev was significantly and inversely associated with age of onset of PD (2/2 studies)^68, 72^, but further analysis within the same clinical cohort (PPMI) using Horvath1 EAA found no association with age of onset^68^.

In longitudinal analyses within a general cohort (n = 149), Hannum AgeDev was significantly associated with increased risk of PD (OR = 1.83, 95% CI 1.08 – 3.13)^73^. Inconsistent evidence were reported for PhenoAge EAA and the risk of PD (1^73^/2 studies)^98^, whereas Horvath AgeDev^73^, Horvath EAA (2/2 studies)^64, 98^, Hannum EAA^64^, Lin EAA^64^, EpiTOC EAA^64^, PhenoAge AgeDev^73^, GrimAge 1 EAA (2/2 studies)^64, 98^, GrimAge 2 EAA^64^, DunedinPoAM^64^ and DundedinPACE (2^64, 73^/3 studies)^98^, AdaptAge^64^ and DamAge^64^ showed no association with risk of PD (Supplementary Figure 8). In longitudinal analysis within clinical populations, Horvath1 AgeDev was negatively associated with the age of onset of PD (2/2 studies), whereas Horvath1 EAA was not, highlighting potential age bias of using AgeDev as a representation of age acceleration that may not reflect a causal relationship with PD.

In MR studies, mixed evidence was observed for a causal association between GrimAge EAA and risk of PD (1^45^/2 studies)^60^ whereas there were no causal associations between Horvath1 IEAA, Hannum EAA and PhenoAge EAA with risk of PD^45^ (eFigure 9). There were no causal effects of PD on Horvath1, Hannum, GrimAge, or PhenoAge^45, 47, 60^.

### Multiple Sclerosis

Five observational studies were in included: four in MS populations (n = 1864), and one general population cohort (Generation Scotland, n =18 821), along with three MR studies using identical EAA GWAS and MS GWAS (n=115 803).

In cross-sectional analyses of MS populations, MS diagnostic status was significantly associated with higher PhenoAge EAA (Epidemiological Investigation of Multiple Sclerosis, EIMS, n=320)^78^, DeepMAge (Gene Expression Omnibus, GEO, n=279)^75^ and AltumAge (GEO, n=39)^76^, and inconsistently associated with GrimAge EAA (1^77^/2 cohorts)^76^. Horvath1 EAA showed no differences between MS cases and controls^76^ (eFigure 10). In longitudinal analyses of a general population cohort (Generation Scotland; n=18 821), no epigenetic clock’s EAA was significantly associated with 10-year risk of incident MS in fully adjusted models accounting for age, sex, cell-type proportions, smoking pack-years, alcohol assumption, social deprivation and education^64^. The strongest signal was for Hannum EAA (HR=1.261, 95% CI 0.99 – 1.61, p = 0.06), which did not reach statistical significance^64^.

In MR studies, Hannum EAA was causally associated with higher risk of MS (2/2 studies)^45, 79^ whereas there were no causal associations between Horvath1, GrimAge or PhenoAge with risk of MS^45, 47, 79^ (Supplementary Figure 11). There were no causal effects of MS on Horvath1 (3/3 studies)^45, 47, 79^, Hannum (2^47, 79^/3 studies)^45^, GrimAge (3/3 studies)^45, 47, 79^, and PhenoAge (2^45, 79^/3 studies)^47^.

### Amyotrophic Lateral Sclerosis

For ALS, three observational studies from ALS populations (n=8064 individuals), and one MR study (ALS GWAS: n=80 610 individuals) were included.

In cross-sectional analyses, GrimAge AgeDev and GrimAge EAA were significantly higher in ALS individuals compared to controls^81^, whereas there were no differences in Horvath EAA, PC-GrimAge EAA, and DunedinPACE between ALS patients and controls^82^. Longitudinal analyses showed that Horvath AgeDev and GrimAge AgeDev were negatively associated with age of onset^80^ and ALS survival^81^ respectively. In MR studies, no causal effects of ALS on Horvath1 EAA, Hannum EAA, PhenoAge EAA, or GrimAge EAA were reported^47^.

### Meta-analysis

Horvath1 EAA (seven studies; nine estimates)^50, 53, 56–58, 64, 65^, Hannum EAA (six studies; eight estimates)^53, 56–58, 64, 65^, PhenoAge EAA (seven studies; nine estimates)^50, 53, 56–58, 64, 65^, GrimAge EAA (seven studies; ten estimates)^50, 53, 56–58, 64, 65^, and DunedinPACE (five studies; six estimates)^50, 53, 58, 64, 65^ were included in the meta-analyses (Fig. 1).

In the minimally adjusted analyses, GrimAge/GrimAge2 EAA was significantly associated with higher risk of all-cause dementia (HR per SD 1·145 [95% CI 1·034–1·269], HR per 1 year 1·039 [95% CI 1·009–1·069], I²=43·7%, p=0·016; Figure 4A), with PhenoAge EAA showing a similar direction, but not statistically significant (1·102 [95% CI 0·999–1·217], I2=48·4%, p=0·052). Restricting to GrimAge EAA, the association remained significant (1·138 [95% CI 1·044–1·241] per SD; 1·037 [1·012–1·062] per 1 year, I²=15·5%, p=0·010). GrimAge2 EAA alone contributed to only two estimates, below the three-estimate threshold for pooling. No significant associations were observed for Horvath1 EAA, Hannum EAA and DunedinPACE. The following covariates were accounted for in fully adjusted analyses: age (n=7), smoking (n=6)^50, 53, 56, 57, 64, 65^, education (n=5)^50, 53, 56, 64, 65^, sex (n=4)^50, 57, 58, 64^, BMI (n=4)^50, 56, 64, 65^, blood-cell composition (n=4)^53, 58, 64, 65^, cardiovascular or metabolic risk factors (n=3)^53, 56, 57^, race (n=2)^56, 65^, and socioeconomic factors (n=2)^53, 64^. . In the fully adjusted models, GrimAge EAA remained significantly associated with higher incident all-cause dementia ((HR per SD 1·109 [95% CI 1·024–1·200], HR per 1 year 1·029 [95% CI 1·007–1·052], I²=0·0%, p=0·018; Figure 4B)). No significant association was found for Horvath1 EAA (1·061 [95% CI 0·939–1·198], I2=70·5%, p=0·293), Hannum EAA (0·961 [95% CI 0·899–1·027], I2=0·0%, p=0·189), PhenoAge EAA (1·083 [95% CI 0·988–1·188], I2=29·3%, p=0·079), or DunedinPACE (1·070 [95% CI 0·916–1·249], I2=55·7%, p=0·314; Figure 5). Restricting to GrimAge EAA, the association remained significant (1·095 [95% CI 1·005–1·194] per SD; 1·026 [1·001–1·051] per 1 year, I²=0·0%, p=0·041).

**Figure 5.**
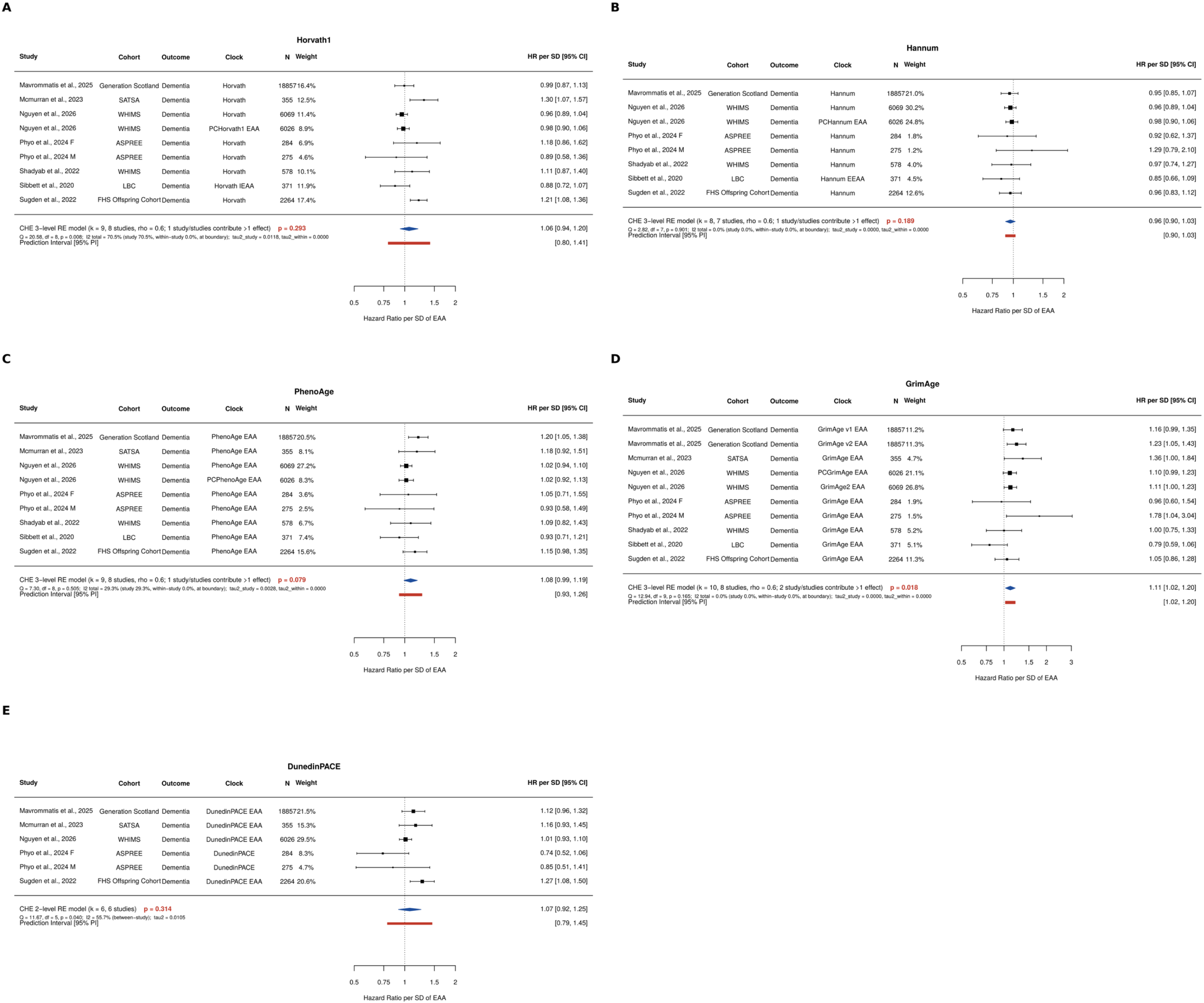
Forest plots of the associations between DNAm EAA or pace of ageing with risk of all-cause dementia across general population cohorts. Note: Forest plots show the pooled hazard ratios (HR) and 95% confidence intervals for the association between the five DNAm clocks and all-cause dementia, derived from random effects meta-analyses. Each row represents an individual study, with the size of the square proportional to the study weight. Female and male results from Phyo et al., 2024 were included as separate entries (denoted F and M) as the study reported sex-stratified estimates. Results are shown for: (A) Horvath1 EAA, (B) Hannum EAA, (C) PhenoAge EAA, (D) GrimAge EAA and (E) DunedinPACE (pace of ageing). To facilitate clinical interpretability, all effect estimates were harmonized to represent HR per one SD higher EAA for Horvath1, Hannum, PhenoAge, and GrimAge and per 0.1-unit higher pace of ageing for DunedinPACE. Note: EAA: epigenetic age acceleration. Heterogeneity was assessed using Cochran’s Q test and the I² statistic.

For GrimAge EAA, between-study heterogeneity was low (I²=0%; Q=11.29, df=7, p = 0.127), and visual inspection of the funnel plot suggested no small-study effects. The pooled estimate was consistent in magnitude across leave-one-out analysis (HR range 1.08-1.12), but its statistical significance depended on the inclusion of individual studies (Supplementary Figure 13-17).

## DISCUSSION

This study reviewed 44 primary studies examining DNAm age across dementia, PD, MS and ALS, providing quantitative synthesis of longitudinal associations between DNAm clocks and incident all-cause dementia, identifying GrimAge EAA as the only clock showing statistically significant associations with dementia risk. Evidence for other neurodegenerative diseases were heterogeneous across clock generations, definitions of epigenetic age acceleration, and study designs. Mendelian randomization analyses provided limited and generally inconsistent evidence for a causal effect of EAA on risk of neurodegenerative diseases, with the only reproducible evidence observed for Hannum EAA and risk of multiple sclerosis.

Whereas previous reviews found no consistent associations between DNAm clocks and dementia owing to study heterogeneity and small individual study samples, our meta-analysis provides empirical support for a prospective association between GrimAge EAA and incident all-cause dementia, supporting its prioritization for further evaluation in dementia risk-prediction models. The pooled association with incident dementia showed low between-study heterogeneity and remained evident across studies adjusting for major dementia risk factors including smoking, BMI, hypertension and socio-economic status^100^, and, where available, leukocyte composition or cell-type proportions. This reduces the likelihood that the association is attributable to conventional vascular risk factors or differences in peripheral blood-cell composition that can influence DNAm age estimates. Compared with first-generation clocks, which may capture a larger contribution from stochastic epigenetic noise, second-generation clocks appear to incorporate more non-stochastic biologically-directed processes relevant to health and disease^101^. GrimAge and GrimAge2 were constructed using DNAm surrogates of smoking exposure and circulating proteins including growth differentiation factor-15 (GDF-15), C-reactive protein (CRP), and hemoglobin A1c (HbA1c), thereby capturing systemic ageing processes that may drive downstream associations with vascular, metabolic, inflammatory, and behavioural risk pathways relevant to dementia. This interpretation is concordant with reported associations between GrimAge EAA and cortical thinning in AD-relevant regions including entorhinal cortex and fusiform gyrus, white matter hyperintensity burden, and other neuroimaging markers of neurodegeneration and cerebrovascular injury^42, 102^. Compared to other epigenetic clocks, GrimAge also showed stronger correlations with brain age acceleration derived from brain morphology measured with MRI^103, 104^. More recent longitudinal data linking baseline epigenetic ageing measures to trajectories of Alzheimer’s disease and related dementias (ADRD) plasma biomarkers including p-tau 181 and p-tau 217 further suggest that DNAm ageing may capture biological vulnerability in amyloid-β and tau pathology that precedes or accompanies disease-specific progression^105^. Besides disease monitoring, PC-GrimAge has also recently demonstrated robust responsiveness across longevity interventions and populations, heralding its value in future research and clinical trials^106^. Our findings position GrimAge as an integrative marker of systemic susceptibility to dementia, whereas its clinical relevance will depend on whether it provides incremental information beyond established plasma biomarkers in determining disease risk, onset and progression, as well as its discriminative ability between ADRDs^107, 108^.

The predominantly null or inconsistent findings for other clocks may partly reflect a mismatch of parameters integrated in DNAm clock training and biological processes driving individual neurodegenerative diseases. First-generation clocks were optimized primarily for chronological age, whereas subsequent generation clocks trained on mortality or longitudinal physiological change may still only be indirectly related to neurodegenerative pathology. Ageing increases vulnerability to neurodegeneration, but the relative contribution of ageing-related processes differs across diseases and may interact with disease-specific genetic, immune and molecular pathways^109^. Emerging ‘explainable’ pathway- and system-informed measures may be more informative in capturing multidimensions of ageing at the level of physiological systems or specific biological processes that may detect disease-related biological variation. In our review, these newer DNAm age was able to differentiate individuals with specific neurodegenerative diseases from controls. For example, PathwayAge was elevated in AD and PD, AltumAge was higher in MS compared to controls, while the Inflammation score showed the most significant difference between AD and controls among the Systems Age scores^13, 76, 110^. The prominence of pathways related to synaptic transmission, neuroimmune regulation, and cellular signaling in the association with AD and PD is biologically plausible given the contribution of peripheral and central immune dysregulation to neurodegeneration^109, 111^. Activation of peripheral adaptive and innate immune cells can amplify central neuroinflammatory processes through disruptions of blood-brain barrier, sustained elevation of pro-inflammatory mediators, excessive microglial phagocytosis and impaired clearance of phosphorylated tau, amyloid-beta (Aβ) or α-synuclein^112–114^. These findings are biologically plausible, but remain predominantly cross-sectional and therefore cannot distinguish antecedent vulnerability from epigenetic remodelling secondary to established disease. Prospective validation of pathology-informed DNAm measures such as those targeintg inflammaging, immune dysregulation or other ageing mechanisms more proximal to neurodegeneration, will also be essential to determine whether enhanced biological interpretability translates to improved risk prediction for specific neurodegenerative diseases^115^.

Tissue and sample specificity may further explain the inconsistent performance for current clocks. Most included studies measured DNAm age in peripheral blood, whereas defining neuropathological processes typically occur within the central nervous system (CNS). A previous systematic review suggests that EAA measured in disease-relevant CNS-based tissues, such as hippocampal tissue in dementia and spinal cord in ALS, may show more consistent associations with neurodegenerative diseases compared to blood-derived EAA^16^. Direct cross-tissue concordance of genome-wide DNAm between peripheral tissues and live human brain within an individual have been found to be locus-specific to CpG sites^80^. Cross-platform differences (e.g. HM450K, EPICv1, EPICv2) can also influence epigenetic age estimates across tissues^81,82^. Differences in DNAm profiles have been reported across varying sources including cerebrospinal fluid, plasma and serum, highlighting the consideration of sample source for reliable insights into tissue pathology^116^. Future clock development should therefore prioritise CpG signatures with demonstrated cross-tissue relevance or derive tissue-informed models anchored to disease-relevant neurobiology, while retaining the practical accessibility and scalability of peripheral sampling^117^, 118.

The paucity of causal signals in the MR literature further suggests that DNAm clocks may not represent direct causal drivers of pathogenesis for neurodegenerative diseases.^10^ The only causal signals reported were between Hannum EAA and risk of late-onset AD and MS, reinforcing the potential role of immune system ageing in susceptibility of neurodegenerative disease risk^109^. However, the absence of broader causal signals across other DNAm clocks highlights an important methodological challenge: confounding of cell-type heterogeneity in existing bulk-tissue DNAm clocks may have obscured disease-relevant causal dynamics^119^. Cell-type deconvolution studies have identified underlying shifts in lymphocyte and neuronal subsets (‘extrinsic’ ageing) to account for 40% and 12% of an epigenetic clock’s accuracy in blood and brain respectively^120^. Yet, conventional *a posteriori* adjustment of DNAm clock values for immune cell fractions does not address cell-type heterogeneity and is uninformative of biological ageing at cell-type-specific resolution^121^. Emerging cell-type specific clocks therefore represent an important methodological advance. Notably, a semi-intrinsic Glia clock in the temporal lobe has demonstrated improved EAA estimates in AD relative to brain-specific and Horvath1 clocks, suggesting that EAA in AD may be more prominent within the glia temporal lobe compartment.^120^ Future studies can investigate immune cell-type or neuron-type specific clocks to better delineate distinct susceptibilities of cell types to age-dependent neurodegeneration, thereby clarifying which cellular ageing processes confer vulnerability to particular neurodegenerative diseases^122, 123^.

Limitations of this review included substantial heterogeneity across studies in DNAm measures, sample characteristics, analytical approaches, and disease outcome definitions, which precluded meta-analysis for neurodegenerative diseases other than dementia. Most studies were conducted in predominantly White populations, limiting generalisability across ancestral and sociocultural groups. This is particularly relevant given emerging evidence that commonly used DNAm clocks, including ensemble and PC-based clocks, appeared to show inconsistent performance across genetically diverse populations^85^. Sex-specific effects were also insufficiently characterised, despite evidence that associations between GrimAge EAA and incident dementia may differ by sex^53^. Future studies may consider how sex-specific biological pathways might affect the performance of DNAm clocks in neurodegenerative diseases^124^. Literature on epigenetic ageing and cerebrovascular disease (CeVD) remains sparse, despite vascular cognitive impairment and dementia being the second most prevalent form of dementia after AD. The use of DNAm age to track co-occurrence or interaction between cerebrovascular disease and AD would be an important avenue for future research^125^. Relatively few studies examined midlife populations (aged 40-59 years), although this period may represent an important window during which critical biological transitions occur before overt neurodegenerative diseases develop^126, 127^. Longitudinal studies beginning in midlife, with repeated DNAm assessment and better representation of diverse ancestral groups, are therefore needed to clarify the temporal sequence linking accelerated biological ageing with brain changes and subsequent functional decline.

## Conclusion

This systematic review and meta-analysis found limited and inconsistent evidence supporting DNAm age clocks as biomarkers of neurodegenerative diseases. Although GrimAge EAA showed a modest association with incident all-cause dementia, other established clocks showed no significant associations with dementia risk, and findings for other neurodegenerative diseases were heterogeneous. Evidence for causality was limited, and the findings do not establish clinical utility or justify implementation for population-level risk stratification. This highlights the need to develop disease-specific DNAm clocks informed by neurodegenerative pathology and relevant cell types. Pathway analyses should investigate the biological processes underlying disease-associated methylation changes, including neuroimmune dysregulation and synaptic dysfunction, and clarify whether peripheral signatures reflect processes within the brain. Disease-specific clocks will require independent longitudinal validation against disease onset and progression, with demonstrated incremental value beyond established risk factors and disease-specific biomarkers before clinical application can be considered.

## Author contributions

Conception and design: Z. K., J. H. T., W. W., A. B. M.; development of methodology: Z. K., J. H. T., W. W., A. B. M.; acquisition of data: Z. K., J. H. T., W. W.; analysis and interpretation of data: Z. K., J. H. T., A. L., W. W., G. J. B., A. B. M.; writing, review and/or revision of the manuscript: Z. K., J. H. T., A. L., W. W., B. T. T. Y., Y. H., N.K., G. J. B., A. B. M.; and study supervision: A. B. M.

## Ethics and Consent to Participate declarations

Not applicable

## Data Availability

The data analysed in this systematic review and meta-analysis were derived from the studies cited in the manuscript. Search strategies and additional methodological details are provided in Supplementary Table 1 and Appendix Texts 1 and 2, included with this preprint.

## Acknowledgements

The authors would like to thank the National University of Singapore senior librarians for assistance with the construction of the search strategy.

## REFERENCES

1. Feigin VL, Vos T, Nichols E, Owolabi MO, Carroll WM, Dichgans M, Deuschl G, Parmar P, Brainin M, Murray C (2020) The global burden of neurological disorders: translating evidence into policy. Lancet Neurol 19:255–265

2. Steinmetz JD, Seeher KM, Schiess N, et al (2024) Global, regional, and national burden of disorders affecting the nervous system, 1990–2021: a systematic analysis for the Global Burden of Disease Study 2021. Lancet Neurol 23:344–381

3. Gadhave DG, Sugandhi VV, Jha SK, Nangare SN, Gupta G, Singh SK, Dua K, Cho H, Hansbro PM, Paudel KR (2024) Neurodegenerative disorders: Mechanisms of degeneration and therapeutic approaches with their clinical relevance. Ageing Res Rev 99:102357

4. Hou Y, Dan X, Babbar M, Wei Y, Hasselbalch SG, Croteau DL, Bohr VA (2019) Ageing as a risk factor for neurodegenerative disease. Nat Rev Neurol 15:565–581

5. Graves JS, Krysko KM, Hua LH, Absinta M, Franklin RJM, Segal BM (2023) Ageing and multiple sclerosis. Lancet Neurol 22:66–77

6. Jylhävä J, Pedersen NL, Hägg S (2017) Biological Age Predictors. eBioMedicine 21:29–36

7. Gonzales MM, Garbarino VR, Pollet E, Palavicini JP, Kellogg DLJ, Kraig E, Orr ME (2022) Biological aging processes underlying cognitive decline and neurodegenerative disease. J Clin Invest. 10.1172/JCI158453

8. Moqri M, Herzog C, Poganik JR, et al (2023) Biomarkers of Aging for the Identification and Evaluation of Longevity Interventions. Cell 186:3758–3775

9. Bell CG, Lowe R, Adams PD, et al (2019) DNA methylation aging clocks: challenges and recommendations. Genome Biol 20:249

10. Conole ELS, Robertson JA, Smith HM, Cox SR, Marioni RE (2025) Epigenetic clocks and DNA methylation biomarkers of brain health and disease. Nat Rev Neurol 21:411–421

11. Belsky DW, Caspi A, Arseneault L, et al (2020) Quantification of the pace of biological aging in humans through a blood test, the DunedinPoAm DNA methylation algorithm. eLife 9:e54870

12. Sehgal R, Markov Y, Qin C, et al (2025) Systems Age: a single blood methylation test to quantify aging heterogeneity across 11 physiological systems. Nat Aging 5:1880–1896

13. Li P, Zhu J, Wang S, et al (2025) Decoding disease–specific ageing mechanisms through pathway-level epigenetic clock: insights from multi-cohort validation. eBioMedicine 118:105829

14. Conole ELS, Robertson JA, Smith HM, Cox SR, Marioni RE (2025) Epigenetic clocks and DNA methylation biomarkers of brain health and disease. Nat Rev Neurol 21:411–421

15. Chervova O, Panteleeva K, Chernysheva E, Widayati TA, Baronik ŽF, Hrbková N, Schneider JL, Bobak M, Beck S, Voloshin V (2024) Breaking new ground on human health and well-being with epigenetic clocks: A systematic review and meta-analysis of epigenetic age acceleration associations. Ageing Res Rev 102:102552

16. Smith RD, Manavi D, Quigley N, Rafati Fard A, Toumbas G, McMurran CE, Kotter MR, Mowforth OD (2026) The evidence for epigenetic age acceleration in neurological disease: A systematic review. Ageing Res Rev 120:103201

17. Yang T, Xiao Y, Cheng Y, Huang J, Wei Q, Li C, Shang H (2023) Epigenetic clocks in neurodegenerative diseases: a systematic review. J Neurol Neurosurg Psychiatry 94:1064–1070

18. Zhou A, Wu Z, Zaw Phyo AZ, Torres D, Vishwanath S, Ryan J (2022) Epigenetic aging as a biomarker of dementia and related outcomes: a systematic review. Epigenomics 14:1125–1138

19. Tay J, Wang W, Maier Andrea B (2025) In: Glob. Epigenetic Age Consort. https://longevityacademy.sg/research/epigenetic-consortium2025.

20. Tay JH, Barros D, Wang W, Wazny VK, Maier AB (2025) Biological age measured by DNA methylation clocks and frailty: a systematic review and meta-analysis. Lancet Healthy Longev. 10.1016/j.lanhl.2025.100773

21. Shan J, Tay JH, Wang W, Tan R, Joshi R, Maier AB, Feng L (2026) Physical activity and biological age measured by DNA methylation clocks: a systematic review and meta-analysis. Lancet Healthy Longev 100835

22. Kriebs A (2026) Systematic review assesses link between physical activity and epigenetic age. Nat Aging 6:1198–1198

23. Simonson MW, Wang W, Tay JH, Hameed S, Min CH, Khan A, Reji A, Maier AB (2026) Biological age measured by DNA methylation clocks and reproductive history: Systematic review and meta-analysis. Mech Ageing Dev 234:112243

24. Simonson MW, Mitra S, Tay JH, Wang W, Maier AB (2026) Clonal haematopoiesis of indeterminate potential and epigenetic age acceleration: Systematic review and meta-analysis. Ageing Res Rev 121:103259

25. Kroemer G, Maier AB, Cuervo AM, et al (2025) From geroscience to precision geromedicine: Understanding and managing aging. Cell 188:2043–2062

26. Stroup DF, Berlin JA, Morton SC, et al (2000) Meta-analysis of Observational Studies in EpidemiologyA Proposal for Reporting. JAMA 283:2008–2012

27. Bocklandt S, Lin W, Sehl ME, Sánchez FJ, Sinsheimer JS, Horvath S, Vilain E (2011) Epigenetic Predictor of Age. PLoS ONE 6:e14821

28. Morandini F, Rechsteiner C, Perez K, Praz V, Lopez Garcia G, Hinte LC, von Meyenn F, Ocampo A (2023) ATAC-clock: An aging clock based on chromatin accessibility. GeroScience 46:1789–1806

29. de Lima Camillo LP, Asif MH, Horvath S, Larschan E, Singh R (2025) Histone mark age of human tissues and cell types. Sci Adv 11:eadk9373

30. Simpson JR (2014) DSM-5 and Neurocognitive Disorders. J Am Acad Psychiatry Law Online 42:159

31. Sachdev P, Kalaria R, O’Brien J, et al (2014) Diagnostic Criteria for Vascular Cognitive Disorders: A VASCOG Statement. Alzheimer Dis Assoc Disord 28:206–218

32. World Health Organization (2019) International statistical classification of diseases and related health problems (11th ed.).

33. American Psychiatric Association (2022) Diagnostic and Statistical Manual of Mental Disorders | Psychiatry Online.

34. Wells GA, Wells G, Shea B, et al (2014) The Newcastle-Ottawa Scale (NOS) for Assessing the Quality of Nonrandomised Studies in Meta-Analyses.

35. Skrivankova VW, Richmond RC, Woolf BAR, et al (2021) Strengthening the Reporting of Observational Studies in Epidemiology Using Mendelian Randomization: The STROBE-MR Statement. JAMA 326:1614–1621

36. Pustejovsky JE, Tipton E (2022) Meta-analysis with Robust Variance Estimation: Expanding the Range of Working Models. Prev Sci 23:425–438

37. Viechtbauer W (2010) Conducting Meta-Analyses in R with the metafor Package. J Stat Softw 36:1–48

38. Cheung MW-L (2014) Modeling dependent effect sizes with three-level meta-analyses: A structural equation modeling approach. Psychol Methods 19:211–229

39. Borenstein M, Higgins JPT, Hedges LV, Rothstein HR (2017) Basics of meta-analysis: I2 is not an absolute measure of heterogeneity. Res Synth Methods 8:5–18

40. Higgins JPT, Thompson SG, Spiegelhalter DJ (2009) A re-evaluation of random-effects meta-analysis. J R Stat Soc Ser A Stat Soc 172:137–159

41. Sterne JAC, Sutton AJ, Ioannidis JPA, et al (2011) Recommendations for examining and interpreting funnel plot asymmetry in meta-analyses of randomised controlled trials. BMJ. 10.1136/bmj.d4002

42. Bonham LW, Sirkis DW, Pang APS, Sugrue LP, Santamaría-García H, Ibáñez AM, Miller BL, Yokoyama JS, Corley MJ (2025) DNA methylation age from peripheral blood predicts progression to Alzheimer’s disease, white matter disease burden, and cortical atrophy. Npj Dement 1:7

43. Coto-Vílchez C, Martínez-Magaña JJ, Mora-Villalobos L, Valerio D, Genis-Mendoza AD, Silverman JM, Nicolini H, Raventós H, Chavarria-Soley G (2023) Genome-wide DNA methylation profiling in nonagenarians suggests an effect of PM20D1 in late onset Alzheimer’s disease. CNS Spectr 28:174–182

44. Degerman S, Josefsson M, Nordin Adolfsson A, Wennstedt S, Landfors M, Haider Z, Pudas S, Hultdin M, Nyberg L, Adolfsson R (2017) Maintained memory in aging is associated with young epigenetic age. Neurobiol Aging 55:167–171

45. Fan J, Liu Q, Liu X, et al (2024) The effect of epigenetic aging on neurodegenerative diseases: a Mendelian randomization study. Front Endocrinol. 10.3389/fendo.2024.1372518

46. Fransquet PD, Lacaze P, Saffery R, Shah RC, Vryer R, Murray A, Woods RL, Ryan J (2021) Accelerated Epigenetic Aging in Peripheral Blood does not Predict Dementia Risk. Curr Alzheimer Res 18:443–451

47. Guo Y, Ma G, Wang Y, Lin T, Hu Y, Zang T (2024) Causal associations and shared genetic etiology of neurodegenerative diseases with epigenetic aging and human longevity. Aging Cell 23:e14271

48. Hao Y, Tian W, Xie B, Fu X, Wang S, Yang Y (2024) The Causal Relationship between Genetically Predicted Biological Aging, Alzheimer’s Disease and Cognitive Function: A Mendelian Randomisation Study. J Prev Alzheimers Dis 11:1826–1833

49. Inkster AM, Duarte-Guterman P, Albert AY, Barha CK, Galea LAM, Robinson WP (2022) Are sex differences in cognitive impairment reflected in epigenetic age acceleration metrics? Neurobiol Aging 109:192–194

50. McMurran CE, Wang Y, Mak JKL, Karlsson IK, Tang B, Ploner A, Pedersen NL, Hägg S (2023) Advanced biological ageing predicts future risk for neurological diagnoses and clinical examination findings. Brain 146:4891–4902

51. Mills JA, Beach SRH, Dogan M, Simons RL, Gibbons FX, Long JD, Philibert R (2019) A Direct Comparison of the Relationship of Epigenetic Aging and Epigenetic Substance Consumption Markers to Mortality in the Framingham Heart Study. Genes 10:51

52. Murthy M, Rizzu P, Heutink P, Mill J, Lashley T, Bettencourt C (2023) Epigenetic Age Acceleration in Frontotemporal Lobar Degeneration: A Comprehensive Analysis in the Blood and Brain. Cells 12:1922

53. Phyo AZZ, Wu Z, Espinoza SE, Murray AM, Fransquet PD, Wrigglesworth J, Woods RL, Ryan J (2024) Epigenetic age acceleration and cognitive performance over time in older adults. Alzheimers Dement Diagn Assess Dis Monit 16:e70010

54. Rivier CA, Szejko N, Renedo D, Clocchiatti-Tuozzo S, Huo S, de Havenon A, Zhao H, Gill TM, Sheth KN, Falcone GJ (2025) Bidirectional relationship between epigenetic age and stroke, dementia, and late-life depression. Nat Commun 16:1261

55. Savin MJ, Wang H, Pei H, et al (2024) Association of a pace of aging epigenetic clock with rate of cognitive decline in the Framingham Heart Study Offspring Cohort. Alzheimers Dement Diagn Assess Dis Monit 16:e70038

56. Shadyab AH, McEvoy LK, Horvath S, et al (2021) Association of Epigenetic Age Acceleration With Incident Mild Cognitive Impairment and Dementia Among Older Women. J Gerontol A Biol Sci Med Sci 77:1239–1244

57. Sibbett RA, Altschul DM, Marioni RE, Deary IJ, Starr JM, Russ TC (2020) DNA methylation-based measures of accelerated biological ageing and the risk of dementia in the oldest-old: a study of the Lothian Birth Cohort 1921. BMC Psychiatry 20:91

58. Sugden K, Caspi A, Elliott ML, et al (2022) Association of Pace of Aging Measured by Blood-Based DNA Methylation With Age-Related Cognitive Impairment and Dementia. Neurology 99:e1402–e1413

59. Thomas A, Ryan CP, Caspi A, Liu Z, Moffitt TE, Sugden K, Zhou J, Belsky DW, Gu Y (2024) Diet, pace of biological aging, and risk of dementia in the Framingham Heart Study. Ann Neurol 95:1069–1079

60. Zhang Z, Liu N, Pan X, Zhang C, Yang Y, Li X, Shao Y (2023) Assessing causal associations between neurodegenerative diseases and neurological tumors with biological aging: a bidirectional Mendelian randomization study. Front Neurosci. 10.3389/fnins.2023.1321246

61. Nichols E, Steinmetz JD, Vollset SE, et al (2022) Estimation of the global prevalence of dementia in 2019 and forecasted prevalence in 2050: an analysis for the Global Burden of Disease Study 2019. Lancet Public Health 7:e105–e125

62. Eissman JM, Ma Y, Qiao M, et al (2026) Biological age acceleration associates with Alzheimer’s disease plasma biomarker levels. Alzheimers Dement 22:e71005

63. Liu H, Yin Z, Chen X, Wang Z (2025) Exploring causal relationships between epigenetic age acceleration and Alzheimer’s disease: a bidirectional Mendelian randomization study. Clin Epigenetics 17:164

64. Mavrommatis C, Belsky DW, Ying K, Moqri M, Campbell A, Richmond A, Gladyshev VN, Chandra T, McCartney DL, Marioni RE (2025) An unbiased comparison of 14 epigenetic clocks in relation to 174 incident disease outcomes. Nat Commun 16:11164

65. Nguyen S, Lu AT, Horvath S, et al (2026) Epigenetic Clocks of Biological Aging and Risk of Incident Mild Cognitive Impairment and Dementia: The Women’s Health Initiative Memory Study. Aging Cell 25:e70424

66. Imahori Y, Qin C, Tang B, Hägg S (2024) Comprehensive analysis of molecular, physiological, and functional biomarkers of aging with neurological diseases using Mendelian randomization. GeroScience 47:2959–2972

67. Engvig A, Kalleberg KT, Westlye LT, Leonardsen EH (2024) Complementary value of molecular, phenotypic, and functional aging biomarkers in dementia prediction. GeroScience 47:2099–2118

68. Gaare JJ, Brügger K, Nido GS, Tzoulis C (2023) DNA Methylation Age Acceleration Is Not Associated with Age of Onset in Parkinson’s Disease. Mov Disord 38:2064–2071

69. Horvath S, Ritz BR (2015) Increased epigenetic age and granulocyte counts in the blood of Parkinson’s disease patients. Aging 7:1130–1142

70. Mei X, Blanchard J, Luellen C, Conboy MJ, Conboy IM (2023) Fail-tests of DNA methylation clocks, and development of a noise barometer for measuring epigenetic pressure of aging and disease. Aging 15:8552–8575

71. Paul KC, Binder AM, Horvath S, Kusters C, Yan Q, Rosario ID, Yu Y, Bronstein J, Ritz B (2021) Accelerated hematopoietic mitotic aging measured by DNA methylation, blood cell lineage, and Parkinson’s disease. BMC Genomics 22:696

72. Tang X, Gonzalez-Latapi P, Marras C, Visanji NP, Yang W, Sato C, Lang AE, Rogaeva E, Zhang M (2022) Epigenetic Clock Acceleration Is Linked to Age at Onset of Parkinson’s Disease. Mov Disord 37:1831–1840

73. Peng X, Flores-Torres MH, Cortese M, Peng C, Hung AY, Schwarzschild M, Ascherio A, Bjornevik K (2026) Epigenetic aging and the risk of Parkinson’s disease. J Neurol Neurosurg Psychiatry 97:385–392

74. Salluzzo MG, Ferraresi F, Marcolungo L, et al (2025) Epigenetic Signatures in an Italian Cohort of Parkinson’s Disease Patients from Sicily. Brain Sci 16:31

75. Galkin F, Mamoshina P, Kochetov K, Sidorenko D, Zhavoronkov A (2021) DeepMAge: A Methylation Aging Clock Developed with Deep Learning. Aging Dis 12:1252–1262

76. Klose D, Needhamsen M, Ringh MV, Hagemann-Jensen M, Jagodic M, Kular L (2023) Smoking affects epigenetic ageing of lung bronchoalveolar lavage cells in Multiple Sclerosis. Mult Scler Relat Disord 79:104991

77. Maltby V, Xavier A, Ewing E, et al (2023) Evaluation of Cell-Specific Epigenetic Age Acceleration in People With Multiple Sclerosis. Neurology. 10.1212/WNL.0000000000207489

78. Theodoropoulou E, Alfredsson L, Piehl F, Marabita F, Jagodic M (2019) Different epigenetic clocks reflect distinct pathophysiological features of multiple sclerosis. Epigenomics 11:1429–1439

79. Liu H, Zhang H, Yin Z, Hou M (2025) Assessment of relationships between epigenetic age acceleration and multiple sclerosis: a bidirectional mendelian randomization study. Epigenetics Chromatin 18:7

80. Zhang M, Tartaglia MC, Moreno D, Sato C, McKeever P, Weichert A, Keith J, Robertson J, Zinman L, Rogaeva E (2017) DNA methylation age-acceleration is associated with disease duration and age at onset in C9orf72 patients. Acta Neuropathol (Berl) 134:271–279

81. Zhao Y, Li X, Wang K, et al (2024) Epigenetic age acceleration is associated with occupational exposures, sex, and survival in amyotrophic lateral sclerosis. eBioMedicine 109:105383

82. Merbaum P, Zwamborn R, Hop P, Sequencing Consortium PMA, van Rheenen W, Veldink J Dissecting epigenetic age acceleration in amyotrophic lateral sclerosis. Epigenomics 17:1153–1161

83. McCartney DL, Min JL, Richmond RC, et al (2021) Genome-wide association studies identify 137 genetic loci for DNA methylation biomarkers of aging. Genome Biol 22:194

84. Horvath S (2013) DNA methylation age of human tissues and cell types. Genome Biol 14:3156

85. Cruz-González S, Okpala O, Gu E, et al (2026) Methylation Clocks Do Not Predict Age or Alzheimer’s Disease Risk Across Genetically Admixed Individuals. 10.7554/elife.105343.2

86. Liu G, Hong C, Huang Y, Guan B, Liu Y, Liu Z, Luo Y (2026) Racial disparities in the association of epigenetic age acceleration with the transition between normal cognition, mild cognitive impairment and dementia. Glob Transit 8:1–9

87. Horvath S, Oshima J, Martin GM, et al (2018) Epigenetic clock for skin and blood cells applied to Hutchinson Gilford Progeria Syndrome and ex vivo studies. Aging 10:1758–1775

88. Hannum G, Guinney J, Zhao L, et al (2013) Genome-wide Methylation Profiles Reveal Quantitative Views of Human Aging Rates. Mol Cell 49:359–367

89. Zhang Y, Wilson R, Heiss J, Breitling LP, Saum K-U, Schöttker B, Holleczek B, Waldenberger M, Peters A, Brenner H (2017) DNA methylation signatures in peripheral blood strongly predict all-cause mortality. Nat Commun 8:14617

90. Levine ME, Lu AT, Quach A, et al (2018) An epigenetic biomarker of aging for lifespan and healthspan. Aging 10:573–591

91. Lu AT, Quach A, Wilson JG, et al (2019) DNA methylation GrimAge strongly predicts lifespan and healthspan. Aging 11:303–327

92. Lu AT, Binder AM, Zhang J, et al (2022) DNA methylation GrimAge version 2. Aging 14:9484–9549

93. Engvig A, Kalleberg KT, Westlye LT, Leonardsen EH (2024) Complementary value of molecular, phenotypic, and functional aging biomarkers in dementia prediction. GeroScience 47:2099–2118

94. Belsky DW, Caspi A, Corcoran DL, et al (2022) DunedinPACE, a DNA methylation biomarker of the pace of aging. eLife 11:e73420

95. Nguyen PK, Zhao D, Okamura T, Chang KH, Wong ND, Yang E (2025) Atherosclerotic Cardiovascular Disease Risk Prediction Models in China, Japan, and Korea. JACC Asia 5:333–349

96. Ying K, Liu H, Tarkhov AE, Sadler MC, Lu AT, Moqri M, Horvath S, Kutalik Z, Shen X, Gladyshev VN (2024) Causality-enriched epigenetic age uncouples damage and adaptation. Nat Aging 4:231–246

97. Senkevich K, Pelletier A, Sato C, Liu L, Keil A, Gan-Or Z, Lang AE, Postuma RB, Rogaeva E (2024) DNA Methylation Age Acceleration as a Potential Biomarker for Early Onset of Rapid Eye Movement Sleep Behavior Disorder. Ann Neurol 95:190–194

98. McMurran CE, Karlsson IK, Mostafaei S, Wang Y, Gerritsen L, Pedersen NL, Hägg S (2025) Brain DNA Methylation Age, Lifestyle Factors and Dementia in the Swedish Twin Registry. Aging Cell 24:e70117

99. Milicic L, Vacher M, Porter T, et al (2022) Comprehensive analysis of epigenetic clocks reveals associations between disproportionate biological ageing and hippocampal volume. GeroScience 44:1807–1823

100. Livingston G, Huntley J, Liu KY, et al (2024) Dementia prevention, intervention, and care: 2024 report of the Lancet standing Commission. The Lancet 404:572–628

101. Tong H, Dwaraka VB, Chen Q, Luo Q, Lasky-Su JA, Smith R, Teschendorff AE (2024) Quantifying the stochastic component of epigenetic aging. Nat Aging 4:886–901

102. Hillary RF, Stevenson AJ, Cox SR, et al (2021) An epigenetic predictor of death captures multi-modal measures of brain health. Mol Psychiatry 26:3806–3816

103. Petersen KJ, Kim S, Canfield P, et al (2026) The GrimAge clock corresponds to brain ageing and social determinants of health in people with HIV. eBioMedicine. 10.1016/j.ebiom.2026.106492

104. Sant’Anna Barbosa Ferreira P, van Dongen J, den Braber A, Boomsma DI, de Geus EJC, van’t Ent D (2025) Epigenetic age acceleration in peripheral blood correlates with brain-MRI age acceleration. Brain 148:2861–2868

105. Zhang B, McEvoy LK, Nguyen S, et al (2025) Epigenetic clocks and longitudinal plasma biomarkers of Alzheimer’s disease. Alzheimers Dement 21:e70983

106. Sehgal R, Borrus D, Armstrong JF, et al (2026) Responsiveness of epigenetic aging biomarkers to longevity interventions in humans. Nat Med 32:3477–3490

107. Grande G, Valletta M, Rizzuto D, et al (2025) Blood-based biomarkers of Alzheimer’s disease and incident dementia in the community. Nat Med 31:2027–2035

108. Buckley RF, Townsend DL, Birkenbihl CJ, et al (2026) Prognostic Value of Blood-Based P-Tau217 Levels for Progression to Cognitive Impairment. JAMA. 10.1001/jama.2026.12556

109. Weiner HL (2025) Immune mechanisms and shared immune targets in neurodegenerative diseases. Nat Rev Neurol 21:67–85

110. Sehgal R, Markov Y, Qin C, et al (2025) Systems Age: a single blood methylation test to quantify aging heterogeneity across 11 physiological systems. Nat Aging 5:1880–1896

111. Berriat F, Lobsiger CS, Boillée S (2023) The contribution of the peripheral immune system to neurodegeneration. Nat Neurosci 26:942–954

112. Zhang Q, Yang G, Luo Y, Jiang L, Chi H, Tian G (2024) Neuroinflammation in Alzheimer’s disease: insights from peripheral immune cells. Immun Ageing 21:38

113. Qin X-Y, Zhang S-P, Cao C, Loh YP, Cheng Y (2016) Aberrations in Peripheral Inflammatory Cytokine Levels in Parkinson Disease: A Systematic Review and Meta-analysis. JAMA Neurol 73:1316–1324

114. Zhang W, Xiao D, Mao Q, Xia H (2023) Role of neuroinflammation in neurodegeneration development. Signal Transduct Target Ther 8:267

115. Franceschi C, Garagnani P, Parini P, Giuliani C, Santoro A (2018) Inflammaging: a new immune–metabolic viewpoint for age-related diseases. Nat Rev Endocrinol 14:576–590

116. Besin V, Humardani FM (2026) Narrative review: translating DNA methylation in dementia. Clin Chim Acta 579:120688

117. Kalyakulina A, Yusipov I, Trukhanov A, Franceschi C, Moskalev A, Ivanchenko M (2025) EpInflammAge: Epigenetic-Inflammatory Clock for Disease-Associated Biological Aging Based on Deep Learning. Int J Mol Sci 26:6284

118. Grodstein F, Lemos B, Yu L, Iatrou A, De Jager PL, Bennett DA (2021) Characteristics of Epigenetic Clocks Across Blood and Brain Tissue in Older Women and Men. Front Neurosci 14:555307

119. Th J, Kf D, Rc S, et al (2022) Functional genomics analysis identifies T and NK cell activation as a driver of epigenetic clock progression. PubMed

120. Tong H, Guo X, Jacques M, Luo Q, Eynon N, Teschendorff AE (2024) Cell-type specific epigenetic clocks to quantify biological age at cell-type resolution. Aging 16:13452–13504

121. Teschendorff AE, Horvath S (2025) Epigenetic ageing clocks: statistical methods and emerging computational challenges. Nat Rev Genet 26:350–368

122. Li W, Zhang Z, Kumar S, et al (2025) Single-cell immune aging clocks reveal inter-individual heterogeneity during infection and vaccination. Nat Aging 5:607–621

123. Gallrein C, Meyer DH, Woitzat Y, Ramirez-Ramirez V, Vuong-Brender T, Kirstein J, Schumacher B (2026) Aging clocks delineate neuron types vulnerable or resilient to neurodegeneration and identify neuroprotective interventions. Nat Aging 1–20

124. Song Z, Feng D, Rahman NA, et al (2026) Sex-specific biological aging clocks across organs and omics. Nat Med 1–12

125. Sachdev PS, Bentvelzen AC, Gustafson D, et al (2026) Vascular Cognitive Impairment and Dementia (VCID): Clinical Features, Neuropathology and Biomarkers. J Am Coll Cardiol 87:52–76

126. Won J, Gogniat MA, Kurazumi T, Nielson KA (2025) Editorial: Midlife brain health: understanding brain aging in middle-age and effects of interventions to prevent neurodegeneration in late life. Front Aging Neurosci. 10.3389/fnagi.2025.1568500

127. Dohm-Hansen S, English JA, Lavelle A, Fitzsimons CP, Lucassen PJ, Nolan YM (2024) The “middle-aging” brain. Trends Neurosci 47:259–272

