## Supplementary Texts 1 and 2 and Supplementary Table 1 for "Biological age measured by DNA methylation clocks and neurodegenerative diseases: A systematic review and meta-analysis"

### Appendix Text 1. Details of method section

**Search strategy and selection criteria**

The search date was set from 1 January 2011 until 15^th^ April 2026 (the first search was conducted on 10th September 2024, the second on 6th June 2025, and an updated search on 15^th^ April 2026). During the first literature search, a large number of relevant studies was identified, making it impractical to summarise all outcomes in a single manuscript. Therefore, in the present paper, the scope was narrowed to two important age-related outcomes, epigenetic age and frailty. Moreover, the initial registration provided examples of age-related diseases and conditions for each physiological system. However, this list was not exhaustive, and as frailty is an age-related condition characterised by dysregulation across multiple physiological systems, it was not explicitly listed under a specific system. To ensure transparency and clarity, the protocol (CRD42024499021) during the latest search (6^th^ June 2025) was updated, explicitly listing frailty as a multisystem outcome and providing more detail on other outcomes.

**Article selection**

The inclusion criteria consisted of peer-reviewed population-based cohort or case-control studies written in English, including adult humans with a mean or median age of 18 years old and above, examining the cross-sectional or longitudinal associations between DNAm clocks and neurodegenerative diseases.

Only articles that included predictors or outcomes specifically related to DNAm clocks, such as DNAm age, DNAm age acceleration (AgeAcce, also known as epigenetic age acceleration (EAA) or relative age acceleration) or DNAm age deviation (AgeDev, also known as absolute age acceleration) measurements, were considered.^8,33^

DNAm age refers to the actual estimate of a DNAm-based clock for a given sample that the clock is applied to. EAA refers to residuals of a linear regression of DNAm age against chronological age. AgeDev refers to the difference between DNAm age and an individual's chronological age. Higher DNAm age reflects higher biological age, whereas higher AgeDev and EAA indicate biological age exceeding chronological age.

DNAm analyses from blood, saliva, buccal cells and skin were included. First-generation clocks consisted of Horvath1, Hannum, YANG, VIDALBRALO, WEIDNER, BOCKLANDT, and GARAGNANI. Second-generation clocks consisted of GrimAge, PCGrimAge, GrimAge2, PhenoAge, PCPhenoAge, adjusted Methylation Age, EpiTOC (mitotic age), Skin & Blood, MPOA, LEVINE, LIN, Zhang, Zhang EN, and Zhang BLUP. Third-generation clocks consisted of DunedinPACE, and newer clocks included SystemsAge. Deep-learning clocks included AltumAge and DeepMAge.

Ascertainment of neurodegenerative diseases (e.g. dementia, PD, MS, and ALS) were based on gold-standard clinical or research diagnostic criteria, or physician diagnosis. Evaluation of dementia and any of its subtype including Alzheimer’s Disease (AD), fronto-temporal dementia (FTD), Lewy Body Dementia (LBD) were based on one of the following: (1) clinically established criteria such as the Diagnostic and Statistical Manual of Mental Disorders 4^th^ Edition (DSM-IV), the International Statistical Classification of Diseases and Related Health Problems 11^th^ Revision (ICD-11 – Chapter 8 Diseases of the Nervous System), and the Alzheimer’s Association workgroup and National Institutes of Health workgroup (NINCDS/ADRDA) criteria, (2) a combination of physician impression and neuropsychological assessment. PD was mainly assessed by movement disorder specialists. MS was measured using the McDonald’s criteria.

**Data extraction**

Estimates including Pearson correlation (*r*), Spearman correlation (ρ), hazards ratio (HR), incident risk ratio (IRR), odds ratio (OR), relative risk ratio (RRR), mean differences, unstandardized and standardized beta coefficients (𝛽s), standard error (SE), 95% confidence interval (95% CI) and its statistical significance (p-value). For MR studies, inverse-weighted variance (IVW) estimates, sensitivity analyses of MR-Egger regression, weighted median and weighted mode, statistical significance (p-value), and multiple testing correction (Bonferroni or Benjamini-Hochberg false discovery rate) were extracted.

**Data synthesis and Meta-analysis**

In observational studies, cross-sectional and longitudinal associations were considered significant at the statistical threshold of p < 0.05. Associations were observed to be showing a trend towards significance when p-value is between 0.05 and 1.0.

In MR studies, a causal relationship was concluded if the direction and estimates of the causal effects of the IVW and sensitivity methods were consistent and the p value with the FDR was less than 0.05 after correction for heterogeneity and horizontal polymorphism. A p < 0.05 but with an FDR > 0.05 was interpreted as a suggestive causal relationship.

*I²* representing the proportion of variability among study outcomes, along with the chi-squared test and its associated p-value, was calculated and presented in the forest plot. *I²* was defined as low (*I^2^* ≤25 %), moderate (*I^2^* >25 % and <75 %), or high (*I^2^* ≥75 %) heterogeneity.^38,40^ χ² (Cochran’s Q test) refers to the observed variability in study estimates exceeds that expected by sampling error alone; a low p-value suggests presence of heterogeneity. ^40^ τ² (Tau-squared) refers to the estimates the absolute between-study variance in effect sizes, with larger values indicating greater heterogeneity; used in random-effects modelling to weight studies.^40^

**Random-effects meta-analyses** were conducted using the *restricted maximum-likelihood estimator (REML)* to pool log-transformed hazard ratios across studies. The **Knapp–Hartung adjustment** was applied to provide robust standard errors and confidence intervals for the pooled estimate. Pooled effects were back-transformed to the HR scale for interpretation.

### Appendix Text 2. Methods for standardised regression coefficient (also known as standardised beta coefficient) (βs) effect size conversions

All effect sizes were standardised into standardised regression coefficients (βs) using established conversion methods adapted from previous study^33, 34, 35.^ The following procedures were used depending on the type of effect size reported in the original studies:

1. From Pearson Correlation Coefficient (r)

- When only correlation coefficients were reported, we used r directly as βs since r equals βs in simple regression (univariate linear regression):
  - βs = r
- Standard error was calculated as:
  - SE(βs) = (1 - r²) / √(n-1),
  - where n is the sample size

2. From unstandardised regression coefficient (β)

- When unstandardised coefficients (β) were reported, we converted to βs using:
  - βs = (SD(X)/SD(Y)) × β
- where SD(X) is the standard deviation of the predictor variable and SD(Y) is the standard deviation of the outcome variable
- Standard error was calculated as:
  - SE(βs) = (SD(X)/SD(Y)) × SE(β)

3. From binary group comparisons

- For studies reporting mean differences between groups:
- We calculated the unstandardised coefficient as
  - β = M₁ - M₂ (where M₁ and M₂ are group means)
- Standard deviation for dichotomous predictors was calculated as:
  - SD(X) = √((n₁n₂)/(n₁+n₂)²)
- The standardised coefficient was then computed as:
  - βs = (SD(X)/SD(Y)) × (β)

Standard error conversion:

- When standard error was not directly reported, we estimated them using:
- From 95% confidence intervals (95% CI):
  - SE = (95%CI upper limit – 95% CI lower limit)/3·92

When SD(X) or SD(Y) is not directly reported:

- From interquartile range (IQR):
  - SD(Y) = IQR/1·349
- From range:
  - SD(Y) = (maximum-minimum)/6
- From standard error:
  - SD(Y) = SE × √n

### Supplemental Table 1. Comprehensive database search terms

| **Database** | **Search Terms, Limits and Filters** | **No. of Searches** |
| --- | --- | --- |
| Embase | ('epigenetic clock*' OR 'epigenetic ag*' OR 'methylation ag*' OR 'dnam clock*' OR 'dnam ag*' OR 'dnam-based biomarker*' OR 'dna methylation clock*' OR 'dna methylation ag*' OR 'dna methylation biomarker' OR 'dna methylation-based biological ageing' OR 'epigenetic-based age acceleration' OR 'epigenetic predictor of age' OR ('biological ag*' AND ('methylation'/exp OR methylation OR methylated)) OR 'horvath* clock' OR 'hannum* clock' OR hannumage OR 'phenotypic age' OR 'phenoageaccel' OR phenoage OR grimage OR dunedinpoam OR dunedinpace OR dnamtl OR 'dnam crp' OR mrscore OR 'mrscore 8cpgs') AND [2011-2026]/py | 7565 |
| Conchrane (CENTRAL) | "epigenetic clock" OR "epigenetic age" OR "methylation age" OR "DNAm clock" OR "DNAm age" OR "DNAm-based biomarker" OR "DNA methylation clock*" OR "DNA methylation age" OR "DNA methylation biomarker" OR "DNA methylation-based biological ageing" OR "Epigenetic-based age acceleration" OR “Epigenetic predictor of age” OR ("biological age" AND (methylation OR methylated)) OR "Horvath clock" OR HorvathAge OR "Hannum clock" OR HannumAge OR "Phenotypic Age" OR "PhenoAgeAccel" OR PhenoAge OR GrimAge OR DunedinPoAm OR DunedinPACE OR DNAmTL OR "DNAm CRP" OR MRscore OR MRscore-8CpGs in Title Abstract Keyword | 250 |
| PubMed | "epigenetic clock*" OR "epigenetic ag*" OR "methylation ag*" OR "DNAm clock*" OR "DNAm ag*" OR "DNAm-based biomarker*" OR "DNA methylation clock*" OR "DNA methylation ag*" OR "DNA methylation biomarker" OR "Horvath clock" OR "Hannum clock" OR HannumAge OR "Phenotypic Age" OR PhenoAgeAccel OR PhenoAge OR GrimAge OR DunedinPoAm OR DunedinPACE OR DNAmTL OR "DNAm CRP" OR MRscore OR MRscore-8CpGs OR OR "DNA methylation"[MeSH Terms] AND "Biomarkers"[MeSH Terms]  Filter:   - From 2011 to 15 April 2026 | 19424 |
| Ovid (PsycINFO and Medline) | "epigenetic clock*" OR "epigenetic ag*" OR "methylation ag*" OR "DNAm clock*" OR "DNAm ag*" OR "DNAm-based biomarker*" OR "DNA methylation clock*" OR "DNA methylation ag*" OR "DNA methylation biomarker" OR "DNA methylation-based biological ageing" OR "Epigenetic-based age acceleration" OR “Epigenetic predictor of age” OR ("biological ag*" AND (methylation OR methylated)) OR "Horvath* clock" OR "Hannum* clock" OR HannumAge OR "Phenotypic Age" OR "PhenoAgeAccel" OR PhenoAge OR GrimAge OR DunedinPoAm OR DunedinPACE OR DNAmTL OR "DNAm CRP" OR MRscore OR MRscore-8CpGs  " Limit used: Humans for species, English for article language and Publication date from 2011 onwards | 3588 |
| Scopus | ("epigenetic clock*" OR "epigenetic ag*" OR "methylation ag*" OR "DNAm clock*" OR "DNAm age" OR "DNAm-based biomarker*" OR "DNA methylation clock*" OR "DNA methylation biomarker" OR "DNA methylation-based biological ageing" OR "Epigenetic-based age acceleration" OR "Epigenetic predictor of age" OR "biological ag*" AND methylation OR methylated OR "Horvath* clock" OR "Hannum* clock" OR hannumage OR "Phenotypic Age" OR "PhenoAgeAccel" OR phenoage OR grimage OR dunedinpoam OR dunedinpace OR dnamtl OR "DNAm CRP" OR dnamclockcortical OR mrscore OR "MRscore-8CpGs") within Article title, Abstract, Keywords  Limit used: Humans for species, English for article language and Publication date from 2011 onwards | 5337 |
| Web of Science | ("epigenetic clock*" OR "epigenetic ag*" OR "methylation ag*" OR "DNAm clock*" OR "DNAm ag*" OR "DNAm-based biomarker*" OR "DNA methylation clock*" OR "DNA methylation ag*" OR "DNA methylation biomarker" OR "Horvath clock" OR "Hannum clock" OR HannumAge OR "Phenotypic Age" OR PhenoAgeAccel OR PhenoAge OR GrimAge OR DunedinPoAm OR DunedinPACE OR DNAmTL OR "DNAm CRP" OR MRscore OR MRscore-8CpGs ) and 2026 or 2025 or 2024 or 2023 or 2020 or 2019 or 2022 or 2021 or 2018 or 2017 or 2016 or 2015 or 2014 or 2013 or 2012 or 2011 (Publication Years) | 5859 |

Note: Date of searches were conducted by 15^th^ April 2026, consisted of three searches.
